# An investigation of the association between characteristics of local crisis care systems and service use in a national survey

**DOI:** 10.1101/2023.03.11.23287106

**Authors:** Antonio Rojas-García, Christian Dalton-Locke, Luke Sheridan Rains, Ceri Dare, Cedric Ginestet, Una Foye, Kathleen Kelly, Sabine Landau, Chris Lynch, Paul McCrone, Shilpa Nairi, Karen Newbigging, Patrick Nyikavaranda, David Osborn, Karen Persaud, Nick Sevdalis, Martin Stefan, Ruth Stuart, Alan Simpson, Sonia Johnson, Brynmor Lloyd-Evans

## Abstract

**Background:** In England, a range of mental health crisis care models and approaches to organising crisis care systems have been implemented, but characteristics associated with their effectiveness are poorly understood.

**Aims:** To i) develop a typology of catchment area mental health crisis care systems and ii) investigate how crisis care service models and system characteristics relate to psychiatric hospital admissions and detentions.

**Methods:** Data about crisis systems were obtained from a 2019 English national survey. Latent class analyses were conducted to identify discernible typologies, and mixed effects negative binomial regression models were fitted to explore associations between crisis care models and admissions and detention rates, obtained from nationally-reported data.

**Results:** No clear typology of catchment area crisis care systems emerged. Regression models suggested that provision of a crisis telephone service within the local crisis system was associated to a 11.6% lower admissions rate and a 15.3% lower detention rate. Provision of a crisis café was associated with a 7.8% lower rate of admissions. The provision of a crisis assessment team separate from the crisis resolution and home treatment service was associated with a 12.8% higher rate of admissions.

**Conclusions:** The configuration of crisis care systems varies considerably in England, but we could notderive a typology which convincingly categorised crisis care systems. Our results suggest that a crisis phone line and a crisis café may be associated with lower rates of admission, but crisis assessment teams, separate from home treatment teams, may not be associate to reductions in hospital admission and detentions.

## Introduction

The development and implementation of mental health services that effectively help to avoid unnecessary acute hospital admissions has been a fundamental objective of researchers and policy makers internationally in recent decades (1, 2). Widespread dissatisfaction with inpatient care and reports that some patients are traumatised or retraumatised by admissions, lack of clarity regarding what is achieved therapeutically and high costs are among the concerns about inpatient care: rising rates of compulsory detention in the UK and elsewhere (2) make the need to seek alternatives still more pressing, as compulsory detention is often associated with coercion and contrasts with the principles of consent and collaboration usually seen as essential to good care (3, 4). Consequently, innovative community crisis care models have emerged in the UK and internationally over several decades to provide alternatives to inpatient care and to improve assessment and triage of mental health crisis (2, 5).

Three community service models are relatively well established in England and internationally, and have a research base suggesting that, if well-implemented, they can be an acceptable and comparably effective alternative to hospital admission for at least some patients in mental health crisis. These are: i) Crisis resolution and home treatment teams, which provide rapid assessment and intensive home treatment usually over a few days or weeks (6); ii) Acute Day Units (ADUs), which provide intensive treatment in settings where patients attend for several hours a day for a mixture of individual and group-based activities and therapeutic interventions (7); and iii) crisis houses, non-hospital residential crisis care services (15–16).

The past decade has seen the further development in the UK of models intended to reduce pressures on emergency and inpatient mental health services for which evidence is largely lacking (8–10). Some newer models, including crisis cafés, street triage teams, and crisis assessment services that are distinct from home treatment services, have proliferated, whilst others such as ADUs have decreased (2). This has led to a substantial heterogeneity in local crisis care systems both in terms of the types of crisis service available and the ways in which the system as a whole is organised. Thus far we know of no research investigating which types of service and which ways of organising the system appear most successful in terms of the key aim of avoiding admissions, especially compulsory detentions.

This study therefore involves two main objectives: i) to determine the types of mental health crisis care systems at local area commissioning level (in 2019 in England these were called Clinical Commissioning Groups (CCG)), and ii) to explore what characteristics of the crisis care systems are associated with mental health hospital admissions and detentions. The second objective was split into two specific objectives focused on a) whether three theoretically-derived variables describing crisis care systems (access, choice and integration) are associated with admissions and detentions, and b) exploring if any data-derived variable for the presence of specific service models and system characteristics are associated with admissions and detentions.

## Methods

The study uses data from a cross-sectional national survey of mental health crisis care systems in England. Managers of CRTs were asked to provide information about services for adults within the local catchment area crisis care system (CCG). Data were collected between April 2019 and December 2019 and referred to the service configuration as provided at the beginning of April 2019. Crisis resolution team (CRT) managers were the preferred survey respondents, as they tend to be familiar with crisis services in their area. Additional respondents were approached where necessary to gain complete data about services within a local area, and respondents and NHS Trust leads were invited to check the collected data for accuracy. Descriptive results from this survey has already been published (2).

### Measures

#### Crisis care system characteristics

Data from the national survey in 2019 were originally collected at the level of the local CRT catchment area (CCG) (2). The national survey included items on the catchment area of the CRT and the CCG which commissioned the service. This information was used to produce variables at the CCG level. Where there were multiple CRTs for a single CCG, the data were combined. Please see supplementary material (Appendix 1) for further detail on how data were combined to produce variables at the CCG level.

The variables from the national survey of crisis care in England that were used to characterise local crisis care systems are presented in Table 1. These binary variables captured information about the presence or absence of service models and about system characteristics across 195 CCGs. The service model variables capture whether various crisis services were present within the catchment area, including as crisis assessment/single point of access services, crisis phone line, psychiatric decision units, crisis cafés, and police and ambulance street triage services. The system characteristics variables provided information about the organisation and delivery of mental health care, including joint management at crisis care team level, shared staffing, voluntary sector involvement, and peer-led services.

**Table 1.** Crisis Care System variables

| **Variable** | **Description** |
| --- | --- |
| *Service models* |  |
| Crisis resolution teams | Multidisciplinary teams that provide rapid assessment and intensive home treatment for a limited period during a crisis |
| Crisis assessment/SPoA services | A service which provides rapid response crisis assessment in a community setting but does not also provide home treatment for acute mental health problems. |
| Crisis phone line | A service which provides crisis response and triage via telephone only, without face-to-face contact (separate from the CRHTT or crisis assessment Team) |
| Psychiatric decision units | A dedicated space (separate from an Accident and Emergency department / psychiatric ward / psychiatric liaison team) in which assessment can be conducted and treatment plans developed for patients in mental health crisis who are accessing emergency services. People may typically stay for up to 24–48h. |
| Triage wards | Inpatient psychiatric wards that admit patients for briefer stays than are usual on acute psychiatric wards. Usually these do not admit people compulsorily detained under mental health legislation, and they work closely with local community crisis services to avoid the need to transfer to an acute psychiatric ward. |
| Crisis cafés | A community-based service which provides out of hours assessment and immediate support for people in mental health crisis, intended to reduce pressure on Accident and Emergency departments. |
| Police street triage services | A team where mental health staff work jointly with police services to arrange appropriate assessment and support for people with mental health problems who come to the attention of the police. |
| Ambulance street triage services | A team where mental health staff work jointly ambulance services to arrange appropriate assessment and support for people with mental health problems who come to the attention of the ambulance service. |
| Crisis houses | A time-limited non-hospital residential service for people in mental health crisis. |
| Acute day units | A non-residential day hospital / crisis day service providing support, activity and therapeutic groups, for people in mental health crisis. |
| Crisis family placements | A service involving families offering short-term crisis foster placements in their family home for people in mental health crisis, supported by local crisis services. |
| *System characteristics* |  |
| Integrated management | Presence of joint management at team level between any crisis services |
| Integrated staffing | Presence of shared staffing between any crisis services |
| Voluntary sector provision | Involvement of voluntary sector organisation in providing any crisis service |
| Peer-led services | Presence of peer-led services in delivering any crisis service |

In addition, three further theoretically derived variables were considered: Access, Choice, and Integration. These variables were constructed from the service models variables and are intended to measure three specific concepts. 1) Access (scores range from 0 to 8) was based on the information about 24/7 access to crisis services, accepting self-referrals, accepting referral from other agencies, and providing a 24/7 CRT response. 2) Choice (scores range from 0 to 6) of crisis service was created by using the information on the availability of crisis cafés, crisis houses, Acute Day Units (ADUs), and crisis family placements. 3) Integration (scores range from 0 to 6) was produced with information on mental health crisis services, acute wards, and emergency services in order to capture integrative process on crisis care. Please see Appendix 1 for detailed definitions of these variables.

#### Outcome variables

In order to complement the information collected in the survey, Public Health England (PHE) Fingertips data were accessed (11) to obtain information on hospital admissions and detention at the CCG level. PHE Fingertips is a publicly available health dataset on service use. The two outcome variables were mental health admission and detention rates. The admissions variable was defined as the number of hospital provider spells in secondary mental health services expressed as a rate per 100,000 person years within the catchment area. The mental health detention rate variable was also expressed per 100,000 person years and defined as the number of people detained under the Mental Health Act (Adult Mental Health and Children and Young People services only). The reporting period for both outcomes variables was between 1^st^ April 2018 and 31^st^ March 2019. As detentions were reported quarterly, three-month totals were added together to a cumulative total for a year. The counts for mental health admissions and detentions were calculated by using the population size by CCG in 2018 (12).

#### Covariates/Putative confounders used in analyses

Potential covariates were also obtained for the period 2018-19 from PHE fingertips. These covariates were selected as variables where there were at least theoretical reasons to expect an influence on crisis care teams and mental health services. The selected covariates were: social deprivation (area deprivation score, adults in employment, adults in stable housing), which has been explore as a risk factor for admission and detention (13) and has been found to be related to provision of crisis alternatives e.g. crisis houses (9); psychiatric morbidity (psychosis prevalence), which has been proposed as a driver of admission and detention rates (14) and may also influence the extent and nature of community crisis care provision; area-level demographic characterstics: some groups in society (age, gender, ethnicity) may be at greater risk of admission, but face barriers to accessing community services, reducing the potential impact of community crisis services. For example, people from minority ethnic groups are at higher risk of psychiatric admission and detention, but have lower use of community services than people from White ethnic groups (15, 16); adults in contact with community services, percentage of service users with a crisis plan, as areas with well-resourced/efficient community services may have lower admission and detention rates (17). The latter two variables were selected as available indicators of the quality and reach of (non-crisis) community mental health services. The covariates were used as putative confounders in exploring relationships between crisis system characteristics and the outcome variables (hospital admission and detention rates).

### Statistical analysis

Descriptive statistics were used to summarise the service models and system characteristics, outcomes, and covariates/confounders. We conducted two separate analyses to address our two objectives.

i. Latent class analysis – typologies of mental health local crisis care systems We used latent class analysis (LCA) to explore whether, based on our national survey data, a typology of crisis care systems at the CCG level could be derived. LCA is based on structural equation modelling methods and aims to classify the binary observations of a study by identifying unobserved latent classes. Several LCAs were conducted for one, two and three classes, in order to identify potential underlying patterns that could help to organise the crisis care systems into distinct categories according to service models and system characteristics. The Akaike’s Information Criteria (AIC) and Bayesian Information Criteria (BIC) were utilised to decide on the number of classes. These indices compared the fit of an LCA model with n classes with that of a model with n+1 classes starting with a single class. Therefore, if the addition of more classes does not improve the fit of the model there would be no evidence to support a more fine-grained categorisation (i.e. categorisations with more classes).
ii. Mixed-effects negative binomial models – Characteristics of the crisis care systems and mental health hospital admissions and detentions Mixed-effects negative binomial models were fitted to study the association between service models and system characteristics with mental health hospital admissions and detentions at CCG level. The two main outcomes (mental health admissions and detentions) were counts from April 2018 to March 2019. Given the initial variables were in the form of rates per 100,000 person-years, counts of mental health admissions and detentions were calculated by multiplying the rates by the CCG resident population and then dividing by 100,000. As the CCGs were clustered within 54 NHS Trusts, a random effect for NHS Trust (random intercept) was included to account for within-trust correlation between CCG measures. In addition, the population size by CCG in 2018 was log-transformed and included as an offset variable in the models. Since the population size in 2018 was not available in four CCGs the rate per 100,000 person-year was used as count and the population size was set at 100,000 for these four CCGs. The service models of triage wards and crisis family placements were excluded from the analysis due to their small numbers within crisis care systems.

First, mixed-effects negative binomial models were fitted to explore the association between each theoretically-derived variable (Access, Choice, Integration) and mental health admissions and detentions, unadjusted and adjusted for covariates (employment, accommodation, prevalence of psychosis, service users with crisis care plans, area deprivation score, contact with mental health services, black and minority ethnic groups, gender and age). Secondly, several mixed-effects negative binomial models were conducted to analyse the relationship between each service models and system characteristics, and mental health admissions and detections, unadjusted and adjusted for covariates. All analyses were performed in Stata 17.

#### Ethics

The 2019 survey (2), commissioned by national policy-makers to understand current service provision, met national guidelines for a service evaluation and therefore did not require review from an ethics committee (18). Survey respondents were sent an information sheet and invitation email, and consented to take part by participating in the survey. We consulted Noclor, the research support service overseeing research governance for several NHS Trusts in North London, to confirm it was appropriate to treat this study as a service evaluation. This paper from our research team reports additional analyses using survey data and publicly available data from NHS Fingertips, for which no additional ethical review is required.

## Results

The summary of the frequency of service models and systems characteristics are presented in Table 2. Those service models most frequently found in CCGs were police street triage (65%), crisis telephone line (63%), and crisis houses (52%). Common system characteristics were integrated staffing (74%) and voluntary sector involvement (66%). There were some missing data for some variables within CCGs, ranging from 16 to 39 responses.

**Table 2.**
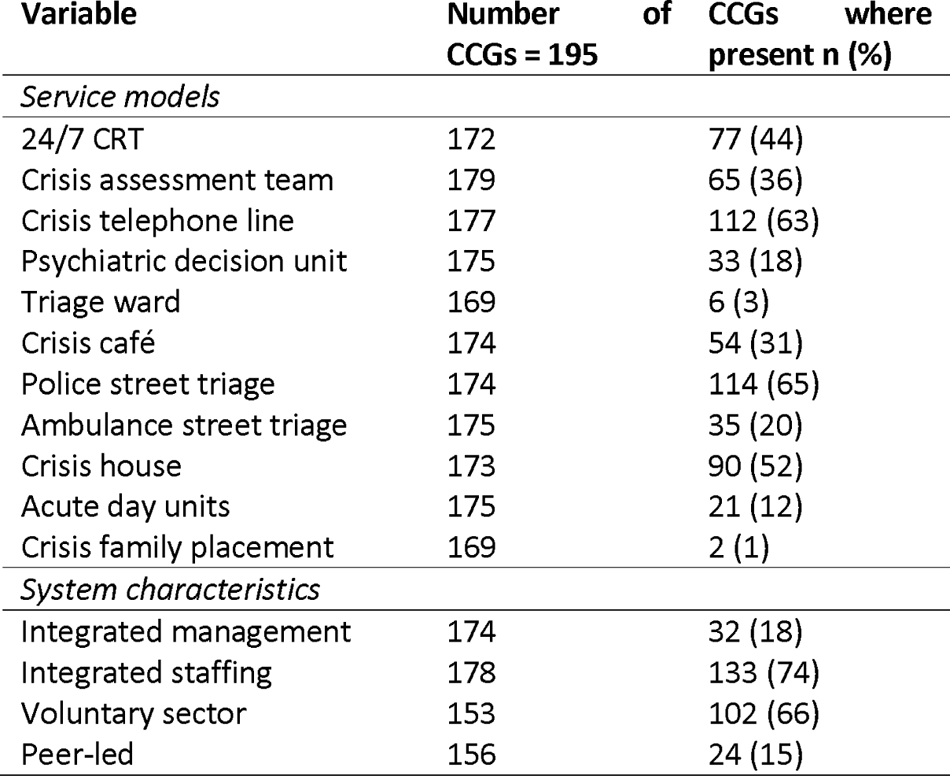
Summary of service model and system characteristics.

Table 3 shows the summary statistics for the outcome variables (mental health admissions and detentions) and for the covariates. In order to provide more explanatory information, median rates of admissions and detentions per 100,000 population are included in the Table. In the analyses, counts (adjusted by population in CCGs) are used.

**Table 3.** Summary of outcomes and covariates

| **Variable** | **N** | **Median** | **IQR** | **Min** | **Max** |
| --- | --- | --- | --- | --- | --- |
| *Outcomes (CCG-level)* |  |  |  |  |  |
| Mental health hospital admissions per 100,000 population (2018/19) | 190 | 666.18 | 557.99 | 59.64 | 800.07 |
| Persons detained under MHA per 100,000 population (2018/19) | 191 | 841.99 | 751.83 | 29.82 | 1135.52 |
|  | **N** | **Mean** | **SD** | **Min** | **Max** |
| *Covariates* |  |  |  |  |  |
| Percentage of CPA adults in employment (2018/19) | 180 | 8.96 | 3.91 | 1.99 | 23.43 |
| Percentage of CPA adults in stable accommodation (2018/19) | 192 | 59.17 | 19.31 | 4.35 | 88.35 |
| Mental health (Psychosis) prevalence (%) all ages (2018/19) | 195 | 0.95 | 0.19 | 0.62 | 1.59 |
| Percentage of service users with crisis plans (2018/19) | 192 | 12.42 | 19.31 | 0.079 | 72.13 |
| Area Deprivation score (IMD 2019) | 189 | 21.92 | 8.03 | 7.18 | 52.13 |
| People in contact with adult mental health services per 100,000 persons (2018/19)* | 195 | 2222.15 | 912.16 | 880.14 | 5488.58 |
| Percentage of population from black and minority ethnic group (2011) | 195 | 13.80 | 15.50 | 1.22 | 72.15 |
| Percentage of population who are male (2017) | 195 | 49.38 | 0.75 | 47.62 | 52.86 |
| Median age of population (2017) | 195 | 40.41 | 4.73 | 27 | 51 |

CCG = Clinical Commissioning Group; CPA = Care Programme Approach; IMD = Index of Multiple Deprivation, IQR = Interquartile range
\* Median and interquartile range

### What types of mental health crisis care system are there, at CCG level?

The first step in the analysis involved trying to obtain a typology of mental health catchment area crisis systems. After conducting several LCA pre-specifying that the analysis should derive two and three different classes, the AIC and BIC suggested that there was no evidence that those models provide a better fit than those with only one class. The models were gradually compared to check how they fitted (i.e. one class vs two classes, two classes vs three classes). The majority of the models did not converge and the models that converged showed large variations within categories. Therefore, the resulting categories did not have a meaningful clinical interpretation in terms of potential typologies for local crisis care systems. These results are presented in the Appendix 2.

### What are the characteristics of crisis care systems associated with inpatient admissions and detentions?

Further planned analysis investigated associations between i) crisis service models available in catchment areas and service use and ii) variables characterising the system as a whole (access, choice and integration) and service use. Regarding the adjusted analyses for crisis service models, it suggested the presence of a crisis telephone line may be associated to an 11.6% lower level of mental health admissions (Incidence Rate Ratio – IRR, 0.884, 95% CI 0.809 to 0.965), while the presence of a crisis café is associated with an 7.8% lower level of in mental health admissions. By contrast, having a crisis assessment team, separate fom to the home treatment team, was associated with a 12.8% higher rate in mental health admissions (IRR, 1.128, 95% CI 1.035 to 1.231). The presence of a crisis telephone line was also associated with a 15.3% lower rate of mental health detentions (IRR, 0.847, 95% CI 0.742 to 0.967). Other service models and all the system characteristics we explored were not associated with admissions and detentions. Our theoretically-derived variables – access, choice, and integration – did not show any association with admissions or detentions (Table 4).

**Table 4.**
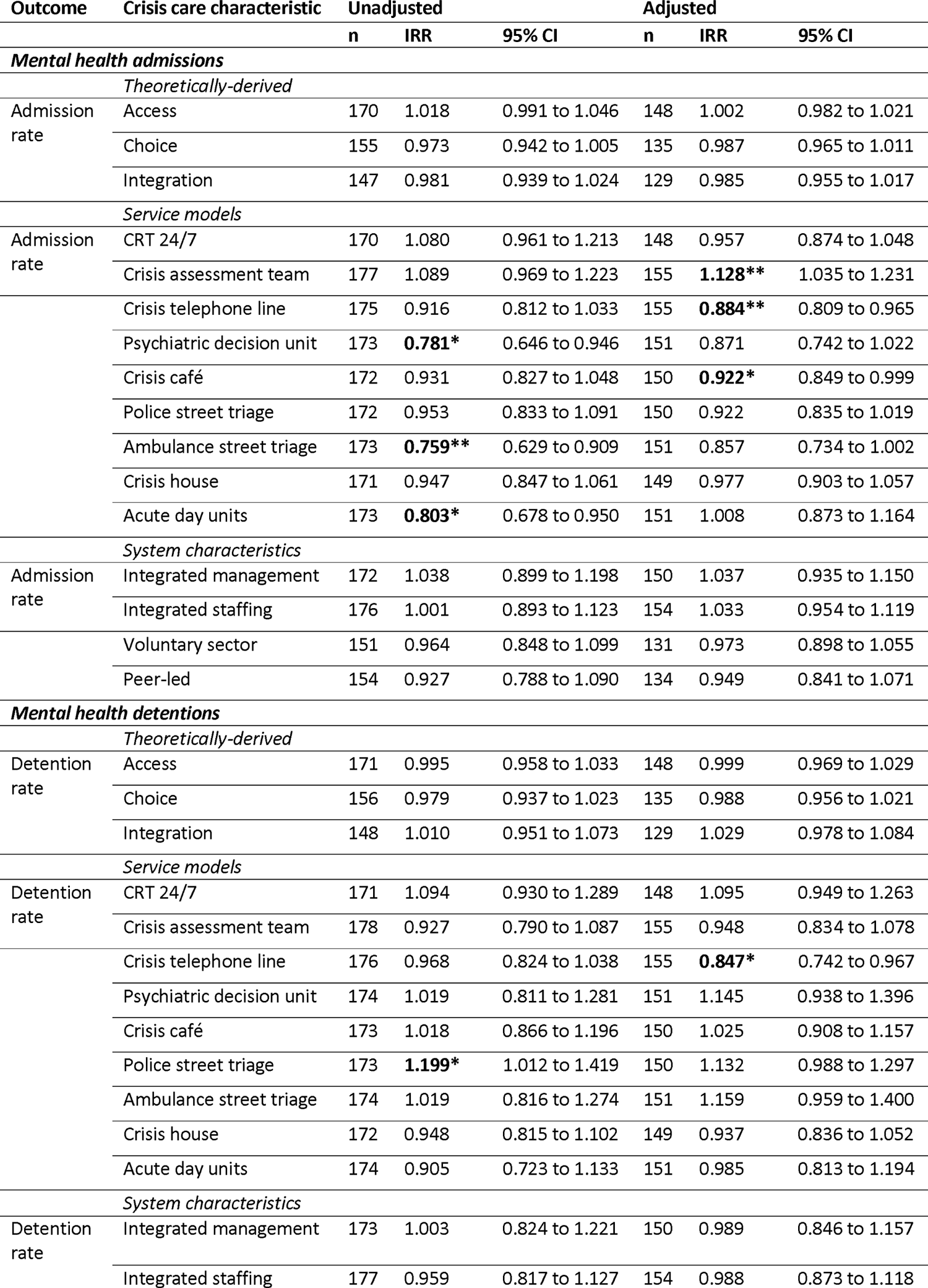
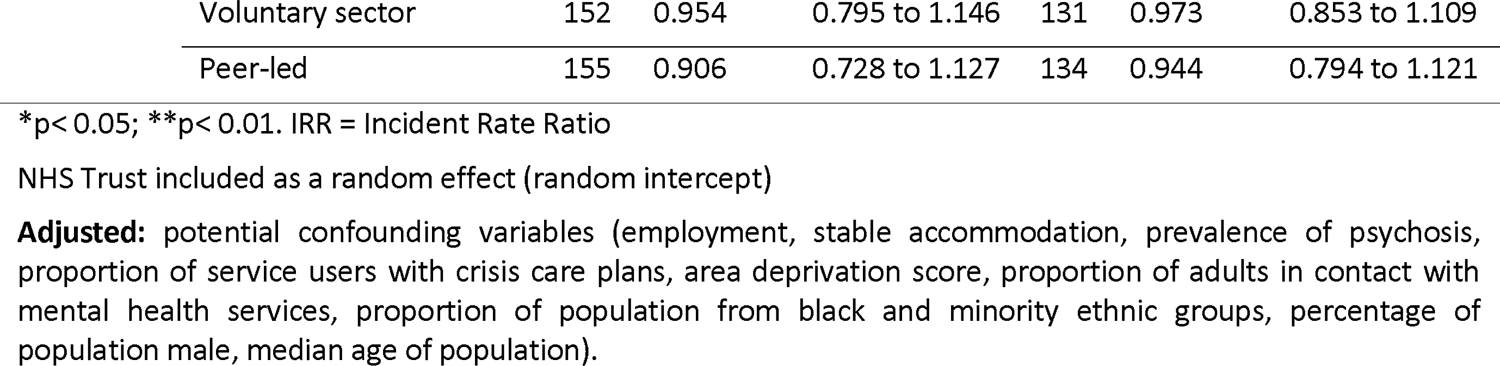
Mixed effects negative binomial models for crisis care characteristics and mental health admissions and detentions

| Outcome | Crisis care characteristic | Unadjusted |  |  | Adjusted |  |  |
| --- | --- | --- | --- | --- | --- | --- | --- |
|  |  | n | IRR | 95% CI | n | IRR | 95% CI |
| ***Mental health admissions*** |  |  |  |  |  |  |  |
|  | *Theoretically-derived* |  |  |  |  |  |  |
| Admission rate | Access | 170 | 1.018 | 0.991 to 1.046 | 148 | 1.002 | 0.982 to 1.021 |
|  | Choice | 155 | 0.973 | 0.942 to 1.005 | 135 | 0.987 | 0.965 to 1.011 |
|  | Integration | 147 | 0.981 | 0.939 to 1.024 | 129 | 0.985 | 0.955 to 1.017 |
|  | *Service models* |  |  |  |  |  |  |
| Admission rate | CRT 24/7 | 170 | 1.080 | 0.961 to 1.213 | 148 | 0.957 | 0.874 to 1.048 |
|  | Crisis assessment team | 177 | 1.089 | 0.969 to 1.223 | 155 | **1.128\*\*** | 1.035 to 1.231 |
|  | Crisis telephone line | 175 | 0.916 | 0.812 to 1.033 | 155 | **0.884\*\*** | 0.809 to 0.965 |
|  | Psychiatric decision unit | 173 | **0.781\*** | 0.646 to 0.946 | 151 | 0.871 | 0.742 to 1.022 |
|  | Crisis café | 172 | 0.931 | 0.827 to 1.048 | 150 | **0.922\*** | 0.849 to 0.999 |
|  | Police street triage | 172 | 0.953 | 0.833 to 1.091 | 150 | 0.922 | 0.835 to 1.019 |
|  | Ambulance street triage | 173 | **0.759\*\*** | 0.629 to 0.909 | 151 | 0.857 | 0.734 to 1.002 |
|  | Crisis house | 171 | 0.947 | 0.847 to 1.061 | 149 | 0.977 | 0.903 to 1.057 |
|  | Acute day units | 173 | **0.803\*** | 0.678 to 0.950 | 151 | 1.008 | 0.873 to 1.164 |
|  | *System characteristics* |  |  |  |  |  |  |
| Admission rate | Integrated management | 172 | 1.038 | 0.899 to 1.198 | 150 | 1.037 | 0.935 to 1.150 |
|  | Integrated staffing | 176 | 1.001 | 0.893 to 1.123 | 154 | 1.033 | 0.954 to 1.119 |
|  | Voluntary sector | 151 | 0.964 | 0.848 to 1.099 | 131 | 0.973 | 0.898 to 1.055 |
|  | Peer-led | 154 | 0.927 | 0.788 to 1.090 | 134 | 0.949 | 0.841 to 1.071 |
| ***Mental health detentions*** |  |  |  |  |  |  |  |
|  | *Theoretically-derived* |  |  |  |  |  |  |
| Detention rate | Access | 171 | 0.995 | 0.958 to 1.033 | 148 | 0.999 | 0.969 to 1.029 |
|  | Choice | 156 | 0.979 | 0.937 to 1.023 | 135 | 0.988 | 0.956 to 1.021 |
|  | Integration | 148 | 1.010 | 0.951 to 1.073 | 129 | 1.029 | 0.978 to 1.084 |
|  | *Service models* |  |  |  |  |  |  |
| Detention rate | CRT 24/7 | 171 | 1.094 | 0.930 to 1.289 | 148 | 1.095 | 0.949 to 1.263 |
|  | Crisis assessment team | 178 | 0.927 | 0.790 to 1.087 | 155 | 0.948 | 0.834 to 1.078 |
|  | Crisis telephone line | 176 | 0.968 | 0.824 to 1.038 | 155 | **0.847\*** | 0.742 to 0.967 |
|  | Psychiatric decision unit | 174 | 1.019 | 0.811 to 1.281 | 151 | 1.145 | 0.938 to 1.396 |
|  | Crisis café | 173 | 1.018 | 0.866 to 1.196 | 150 | 1.025 | 0.908 to 1.157 |
|  | Police street triage | 173 | **1.199\*** | 1.012 to 1.419 | 150 | 1.132 | 0.988 to 1.297 |
|  | Ambulance street triage | 174 | 1.019 | 0.816 to 1.274 | 151 | 1.159 | 0.959 to 1.400 |
|  | Crisis house | 172 | 0.948 | 0.815 to 1.102 | 149 | 0.937 | 0.836 to 1.052 |
|  | Acute day units | 174 | 0.905 | 0.723 to 1.133 | 151 | 0.985 | 0.813 to 1.194 |
|  | *System characteristics* |  |  |  |  |  |  |
| Detention rate | Integrated management | 173 | 1.003 | 0.824 to 1.221 | 150 | 0.989 | 0.846 to 1.157 |
|  | Integrated staffing | 177 | 0.959 | 0.817 to 1.127 | 154 | 0.988 | 0.873 to 1.118 |

|  |  |  |  |  |  |  |
| --- | --- | --- | --- | --- | --- | --- |
| Voluntary sector | 152 | 0.954 | 0.795 to 1.146 | 131 | 0.973 | 0.853 to 1.109 |
| Peer-led | 155 | 0.906 | 0.728 to 1.127 | 134 | 0.944 | 0.794 to 1.121 |

\*p<0.05; \*\*p<0.01. IRR = Incident Rate Ratio
NHS Trust included as a random effect (random intercept)
**Adjusted:** potential confounding variables (employment, stable accommodation, prevalence of psychosis, proportion of service users with crisis care plans, area deprivation score, proportion of adults in contact with mental health services, proportion of population from black and minority ethnic groups, percentage of population male, median age of population).

## Discussion

The study provides exploratory findings on the association of several mental health crisis service models with mental health admissions and detentions. The presence of certain models or characteristics seemed to be related to decreasing fewer admissions s (e.g. crisis café, crisis telephone line) and detentions (e.g. crisis telephone line), whilst only having a separate crisis assessment team was apparently associated with an higher level of mental health admissions. The variables capturing the concepts of access, integration and choice were not associated with either mental health admissions or detentions. Furthermore, the analyses exploring potential categories of crisis care systems did not yield any valid typology.

To the best of our knowledge, this is the first study that analyses the components of crisis care systems to explore whether there are the typologies of such systems, and to explore associations of system characteristics with mental health admissions and detentions. The findings from the adjusted models indicate that the presence of crisis telephone lines, such as a single point of access providing telephone triage, is associated with a lower rate in both admissions and detentions. According to a Care Quality Commission report (19), individuals experiencing a mental health crisis may benefit from crisis telephone services. However, to ensure effective delivery, it is essential to provide a service which is sensitive to-individuals and accessible out-of-hours (19). Similarly, the results of the adjusted analyses indicate that the presence of a crisis café was associated with reduced mental health admissions, but not rates of detention. Crisis cafés are alternative services to hospital emergency departments, usually provided by the voluntary sector, where individuals experiencing a crisis may obtain support and signposting to other services (2). As this is a cross-sectional study, the causal relationships here are unclear. However, such a finding would be in keeping with findings that assessment in emergency departments is disproporationately likely to lead to admission (20), suggesting that providing a potentially more acceptable and accessible alternative to attending emergency departments and diverting people from this setting may be productive in reducing admissions. This finding suggests that access to a crisis café may help avoid admission. However, further investigations of longitudinal nature are needed to confirm this possibility.

Initial local evaluations of early-adopter crisis assessment teams were promising, suggesting that teams focused only on crisis assessment and not on home treatment might prevent admissions more effectively than combined crisis assessment and home treatment teams (21). Nevertheless, we found that nationally, the presence of a stand-alone crisis assessment team in the local crisis system was associated with a higher rate of mental health admissions. Our research cannot elucidate the reasons for this cross-sectional finding. A possible explanation is that separating out initial crisis assessment and crisis home treatment into separate teams and removing the “gatekeeping” of hospital admissions from the service which provides crisis home treatment, may lead to discontinuities in care and more conservative risk management. Crisis Resolution Teams, which provide both crisis assessment and home treatment, could be best placed to limit the access to hospital admissions where they are avoidable.

Triage through a Crisis assessment team could in some contexts act as a barrier to prompt access to CRT support, which might also lead to an increase in admissions. Previous research has also suggested that longer opening hours (22) and better access to CRTs (1) were associated with lower admissions. However, our results did not show any significant association between access, or having a 24-hour CRT, and lower rates of mental health admissions. Furthermore, other theoretically-derived variables such as integration and choice, did not show any association with either admissions or detentions. Likewise, other service models and system characteristics did not show any association with admissions and detentions.

The variables capturing the concepts of access, integration, and choice were not associated with either outcome. This was counter to our expectations, as the variables were developed by a stakeholder group to capture aspects of crisis care system functioning reflecting good practice. Lack of association might reflect imprecision in data collection (one informant often provided information on a whole system), or lack of granularity in recording variables intended to capture the functioning of a whole system.

### Limitations

First, the survey data was self-reported, which could have led to some inaccuracies. PHE Fingertips data is likewise based on locally submitted data, and therefore some biases may be introduced if data was not consistently collected. The three theoretically-derived variables: choice, integration, and access were not validated, and not checked for internal consistency, which may undermine their reliability and validity. Our study only explored cross-sectional relationships between variables and one period of time. The mixed-effect negative binomial regression models were fitted to test association rather than proving causation. In this regard, we were able to adjust for some potentially important covariates, but we cannot ensure that other covariates not included in the analysis may influence the results. Also, it was not feasible to use some originally proposed outcome variables (e.g. A&E attendance) given the poor quality of the data (based on advice from NHS England). In addition, the analyses were not adjusted for multiple testing, which may increase the risk that some significant differences were due to chance. Some corrections could have been applied (e.g., Bonferroni correction). However, the main objective of the study was to explore the relationship between crisis care system characteristics and admissions and detentions for future investigations, rather than to try to establish any impact on the association between variables. Another limitation is that data were collected before the COVID-19 outbreak and many crisis care systems have been reorganised since then. Local commissioning areas in England have also been reorganised since 2019, with CCGs replaced by larger area-level Integrated Care Boards (23)

### Implications for policy, practice and research

Given that our findings were exploratory, the conclusions are preliminary. However, our study does not provide evidence to support any policy recommendation for creating new crisis assessment teams, splitting the crisis assessment and crisis home treatment functions into different teams. Conversely, the positive results for a crisis telephone lines suggest that 24-hour access to crisis support may be important. Our study also provides preliminary evidence that introducing crisis cafes into local crisis care systems could potentially help to reduce admission rates. Overall, the analyses yielded few associations between what service types are included within a local crisis care system and local admissions and detentions. Investing time and money in setting up innovative new crisis services has an opportunity cost. Rather than invest in a proliferation of new crisis service models, local commissioners and service planners may be better advised to focus energies and resources on improving the quality of care in their current local crisis system, which may also help to reduce inpatient admissions (24).

Further research is needed to clarify these exploratory findings. More robust methodological approaches such as longitudinal designs (i.e., time series of controlled studies) may help to reveal the association between crisis care system characteristics and mental health admissions and detentions. Additionally, this research could explore the specification of models for innovative crisis care services and development and evaluation of quality improvement programmes, such as the evaluation of crisis cafes. Other approaches investigating the psychiatric admissions and detentions, and the implementation of certain models may also help to clarify the direction of potential associations.

### Lived Experience Commentary

#### Tamar Jeynes and Lizzie Mitchell

We welcome this exploration into different models of crisis care – a much needed investigation.

Preventing admissions is a crucial part of crisis care, but it should not be the sole focus. Measuring only admission rates does not capture the other benefits crisis care should bring, such as having human connection, helping people to manage extreme levels of distress and preventing harm to themselves, others or dying by suicide. We understand this is hard to measure, but the true effect of crisis services cannot just be judged by admissions – it needs to be what service users feel helps them at their time of need.

Statistical methods provide some insight into what may work. However, the variance of the services cannot be captured. As an example, crisis houses and cafes can be NHS, Third Sector or Peer-run, each using different approaches which may at times contradict the approach of another using the same name.

The finding that crisis telephone lines reduce admissions is of interest because of the wide variation of efficacy between different areas. We wondered if exceptional areas masked the failings of other areas.

Mental health development begins in childhood, yet children and young people’s (CYP) services are not included in this research. To change the way services are structured, we need to change this. Children and young people’s crisis services are often structured very differently to adult services. These should be included so we can see crisis care across a continuum and assess effective structure of services.

The data for this study was collected pre-covid in 2019. The pandemic has led to increased funding into crisis healthcare including research co-produced with lived experience researchers which readers could compare for changes and supporting evidence of findings. Further exploration into such research should be a priority to ensure services are improved for people at the time when they need them the most.

The findings themselves are of interest beyond the scientific community and we welcome MHPRU plans to make plain English summary accessible to lay people.

## Declaration of interests

NS is the director of the London Safety and Training Solutions Ltd, which offers training in patient safety, implementation solutions and human factors to healthcare organisations and the pharmaceutical industry. The other authors have no conflicts of interest to declare.

## Funding

This paper presents independent research commissioned and funded by the National Institute for Health Research (NIHR) Policy Research Programme, conducted by the NIHR Policy Research Unit (PRU) in Mental Health. NS’ research is supported by the National Institute for Health Research (NIHR) Applied Research Collaboration (ARC) South London at King’s College Hospital NHS Foundation Trust. NS is a member of King’s Improvement Science, which offers co-funding to the NIHR ARC South London and is funded by King’s Health Partners (Guy’s and St Thomas’ NHS Foundation Trust, King’s College Hospital NHS Foundation Trust, King’s College London and South London and Maudsley NHS Foundation Trust), and Guy’s and St Thomas’ Foundation. The views expressed are those of the authors and not necessarily those of the NIHR, the Department of Health and Social Care or its arm’s length bodies, or other government departments.

## Data Availability

All data produced in the present study are available upon reasonable request to the authors

## Acknowledgement

Thank you to Lizzie Mitchell and Tamar Jeynes, two members of the NIHR Mental Health Policy Research Unit Lived Experience Working Group, who wrote a Lived experience Commentary to accompany this paper, providing an unmediated reflection on the paper from the perspective of two researchers with first-hand experience of mental distress and using mental health services.

## Author contributions

All authors contributed to designing the study and revising the manuscript. All authors approved the final version. AR-G led data analysis, supported by CD-L and LS-R, and with guidance from statisticians SL and CG. AR-G drafted the manuscript with help from BLE, and subsequent review from all authors. SJ and BLE managed the project.

## Data availability

De-identified data are available from the authors on reasonable request.

## Appendix 1 Access, Choice, Integration (theoretically derived variables)

### Scoring rules

Note: Each variable is first scored at the CRT-level before being transformed to the CCG-level according to the following rules.

1. When coded at the CRT-level, variables were scored as missing where insufficient data were available to establish whether a score of 0, 1 or 2 was achieved
2. All variables score zero if they do not score one or two and are not missing
3. The Domain total = missing if any of the variables scoring on to the domain are missing
4. At the CCG-level score each variable if data are available for at least 50% of CRTs serving this CCG – otherwise count as missing
5. Each criterion as met if at least 50% of CRTs which provided data for this CCG meet the criterion

**Table 1:** Domain 1: Access (Total score 0-8)

| **Table 1: Domain 1: Access (Total score 0-8)** |  |  |  |
| --- | --- | --- | --- |
| **Variable** | **Scoring** | **Rationale: why is this important to people in crisis?** | **Score at CCG-level (N=195)** |
| 24/7 access | 2: There is a Crisis Assessment Team which is open 24/7 OR There is a CRT or Crisis line which offers telephone support 24/7 and the CRT sees people in person 24/7 (including home visits or seeing patients on NHS premises)<br>1: There is a crisis line or CRT offering telephone assessment and support which runs 24/7 but no Crisis assessment Service or CRT which can see people in person 24/7 | Services are running whatever time I need to access crisis support | 0 = 13 (7%)<br>1 = 34 (19%)<br>2 = 129 (73%)<br>Missing = 19 (10%) |
| Self-referral | 2: People known and not known to services can self-refer to the Crisis Assessment Team or CRT<br>1: People known to services can self-refer to the Crisis Assessment Team or CRT | I can access crisis services directly when I need to | 0 = 58 (33%)<br>1 = 33 (19%)<br>2 = 83 (48%)<br>Missing = 21 (11%) |
| Referral from other agencies | One point each for:<br>1: The Crisis Assessment Team or CRT accepts referrals from emergency services (police and/or ambulance)<br>1: The Crisis Assessment Team or CRT accepts referrals from GPs | Other professionals trying to help me can access crisis services directly when they need to | 0 = 28 (16%)<br>1 = 29 (17%)<br>2 = 117 (67%)<br>Missing = 21 (11%) |
| 24/7 CRT response | 2: The CRT can see current service users at home 24/7<br>1: The CRT can see current service users on NHS premises 24/7 | The Crisis Team who are supporting me can see me conveniently whatever time I need help | 0 = 76 (39%)<br>1 = 29 (15%)<br>2 = 84 (43%)<br>Missing = 6 (3%) |

**Table 2:** Domain 2: Choice (Total score 0-6)

| **Table 2: Domain 2: Choice (Total score 0-6)** |  |  |  |
| --- | --- | --- | --- |
| **Variable** | **Scoring** | **Rationale: why is this important to people in crisis?** | **Score at CCG-level (N=195)** |
| Alternative places to seek help in a crisis<br>(Crisis cafes) | 2: There is an immediately accessible place other than A&E where people can go in a mental health crisis (e.g. a crisis café with drop-in or self-referral)<br>1: There is an alternative to A&E e.g. a crisis café, but access is via referral from professionals | I don't have to go to A&E if I feel unsafe at home: there is an alternative | 0 = 119 (69%)<br>1 = 10 (6%)<br>2 = 44 (25%)<br>Missing = 22 (11%) |
| Alternatives to treatment at home in a crisis | 2: There are at least two different crisis alternatives to home treatment or hospital (crisis house, Acute day Unit, family placement scheme)<br>1: There is at least one crisis alternative to hospital or home treatment | There are other options not just home treatment, if I need another environment to support my recovery | 0 = 76 (45%)<br>1 = 78 (46%)<br>2 = 14 (8%)<br>Missing = 27 (14%) |
| Availability of alternatives to treatment at home in a crisis<br>(ADUs, crisis houses or crisis family placements) | One point each for:<br>1: The CRT reports at least one crisis alternative usually has spaces available when required<br>1: Admission to at least one crisis alternative can usually be arranged within 24 hours | And other options in addition to home treatment are usually available | 0 = 76 (47%)<br>1 = 9 (6%)<br>2 = 76 (47%)<br>Missing = 34 (17%) |
| *Types of support | One point each for:<br>1: The local crisis system includes at least one non-NHS provided service (from survey responses or Karen Newbigging project list)<br>1: At least one crisis service is service user-led | If I have not found typical NHS healthcare helpful, other types of support are offered | - |
| *Dropped due to high missing rate |  |  |  |

**Table 3:** Domain 3: Integration (Total score 0-6)

| **Table 3: Domain 3: Integration (Total score 0-6)** |  |  |  |
| --- | --- | --- | --- |
| **Variable** | **Scoring** | **Rationale: why is this important to people in crisis?** | **Score at CCG-level (N=195)** |
| Within mental health crisis services | 2: CRHTT care can be accessed via a single assessment (e.g. the CRHTT accepts self-referrals from all; or the Crisis Assessment Team accepts self-referrals from all and can arrange CRHTT care without reassessment)<br>1: There are two steps to CRHTT care in some or all circumstances (e.g. the CRHTT accepts referrals directly from all health professionals but not self-referrals from all; OR referrals are via the Crisis Assessment Team, which accepts self-referrals, but the CRHTT re-assesses referrals from the Crisis Assessment Team before accepting people for home treatment) | I can get crisis support without having to tell my story repeatedly to different professionals | 0 = 79 (45%)<br>1 = 24 (14%)<br>2 = 74 (43%)<br>Missing = 18 (9%) |
| With acute wards | 2: The CRHTT (or Crisis Assessment Team) report that they seek to gatekeep at least 95% of people in person before admission to an acute inpatient ward<br>1: The CRHTT report that they seek to gatekeep at least 50% of people in person. | I will not be admitted to hospital unless home treatment and crisis alternatives have been fully considered | 0 = 18 (11%)<br>1 = 55 (33%)<br>2 = 92 (56%)<br>Missing = 30 (15%) |
| With emergency services | One point each for:<br>1: There is a police street triage team<br>1: There is an ambulance street triage team | If I come into contact with the police or ambulance staff during a crisis, they can access help for me quickly, in the least restrictive way. | 0 = 58 (36%)<br>1 = 81 (50%)<br>2 = 24 (15%)<br>Missing = 32 (16%) |

## Appendix 2 Summary of Latent Class Analysis for Crisis Care Teams Project CRT 24/7; Crisis assessment; Crisis café; Crisis house (Missing= 29)

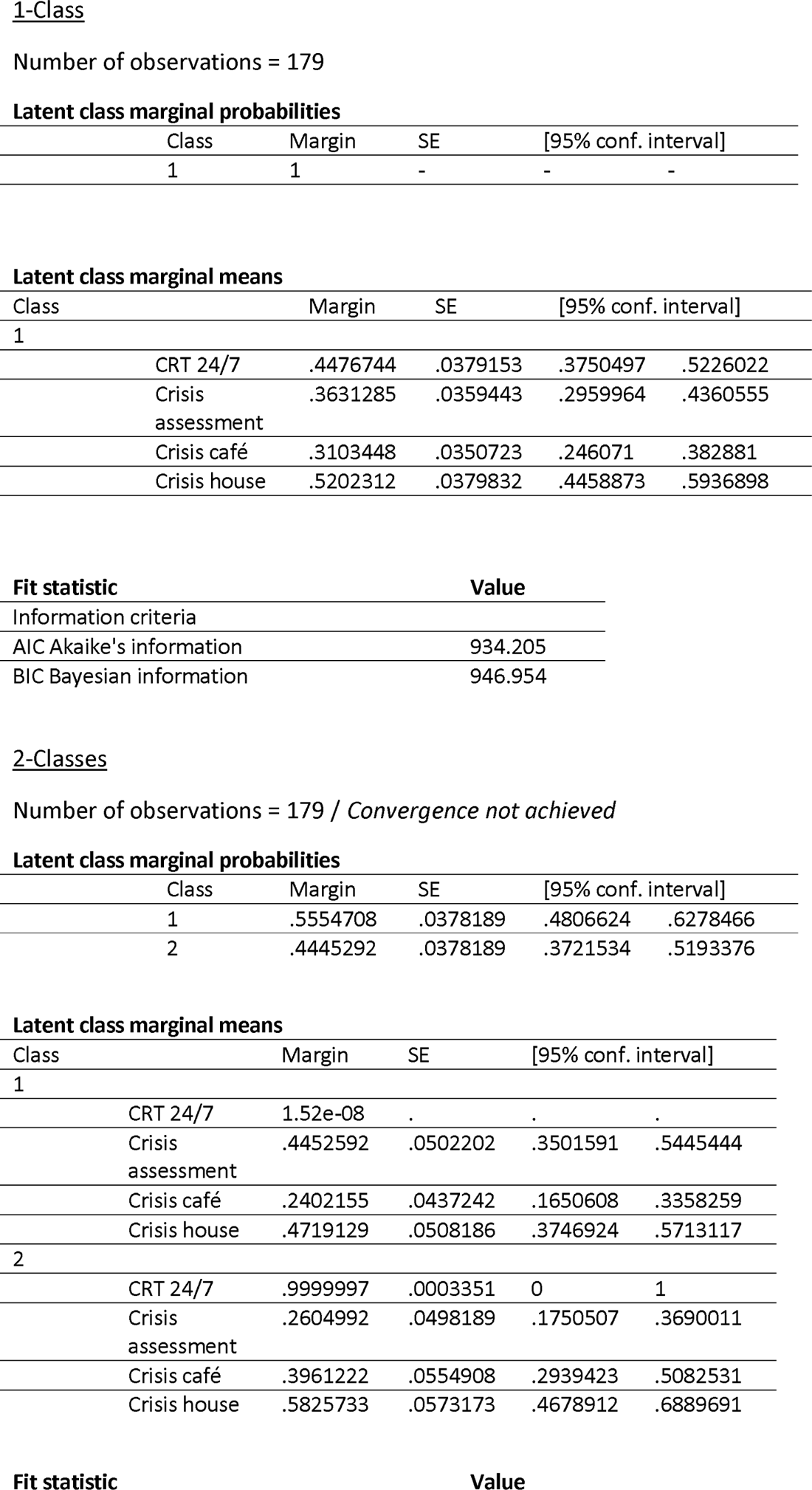
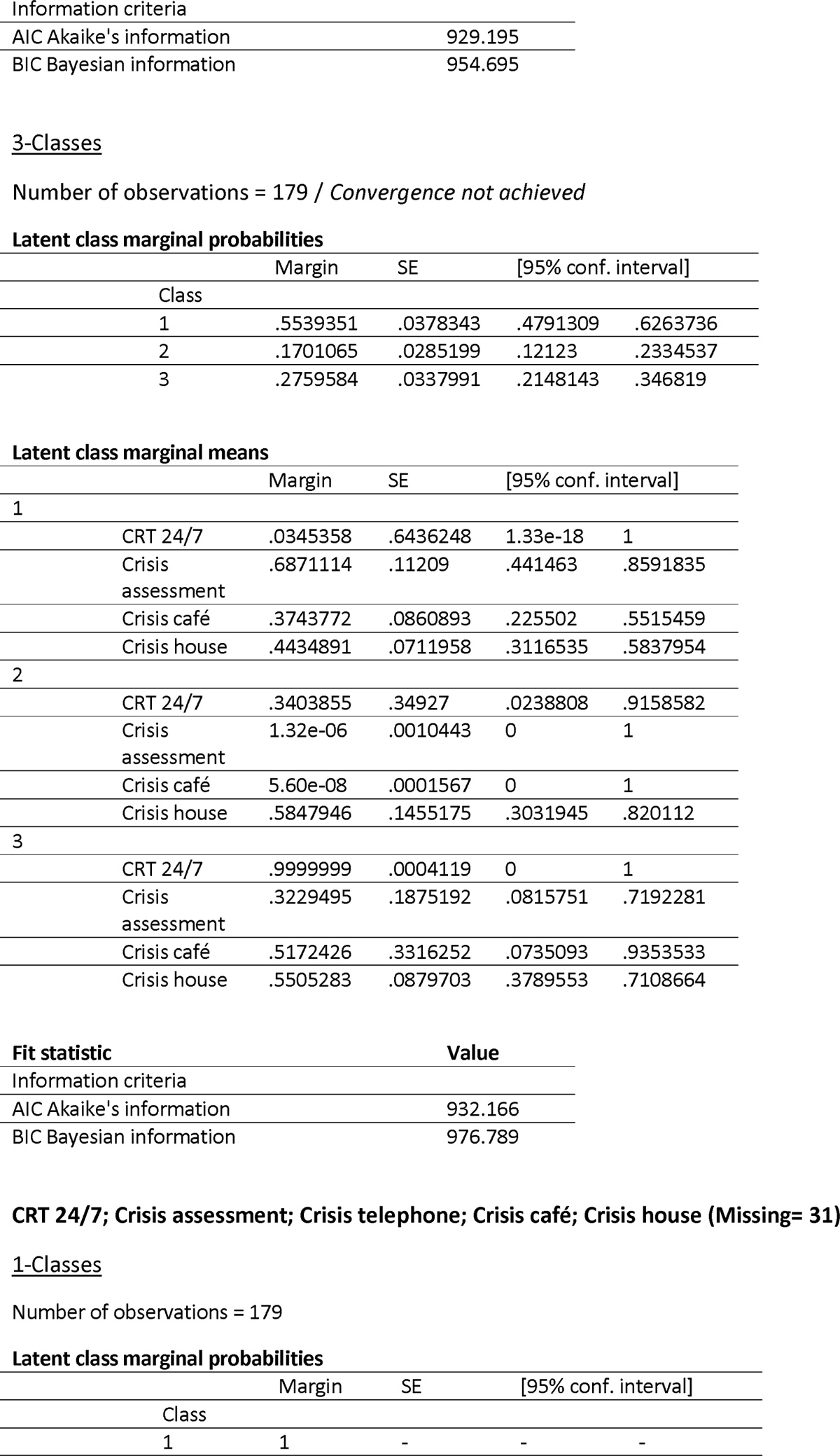
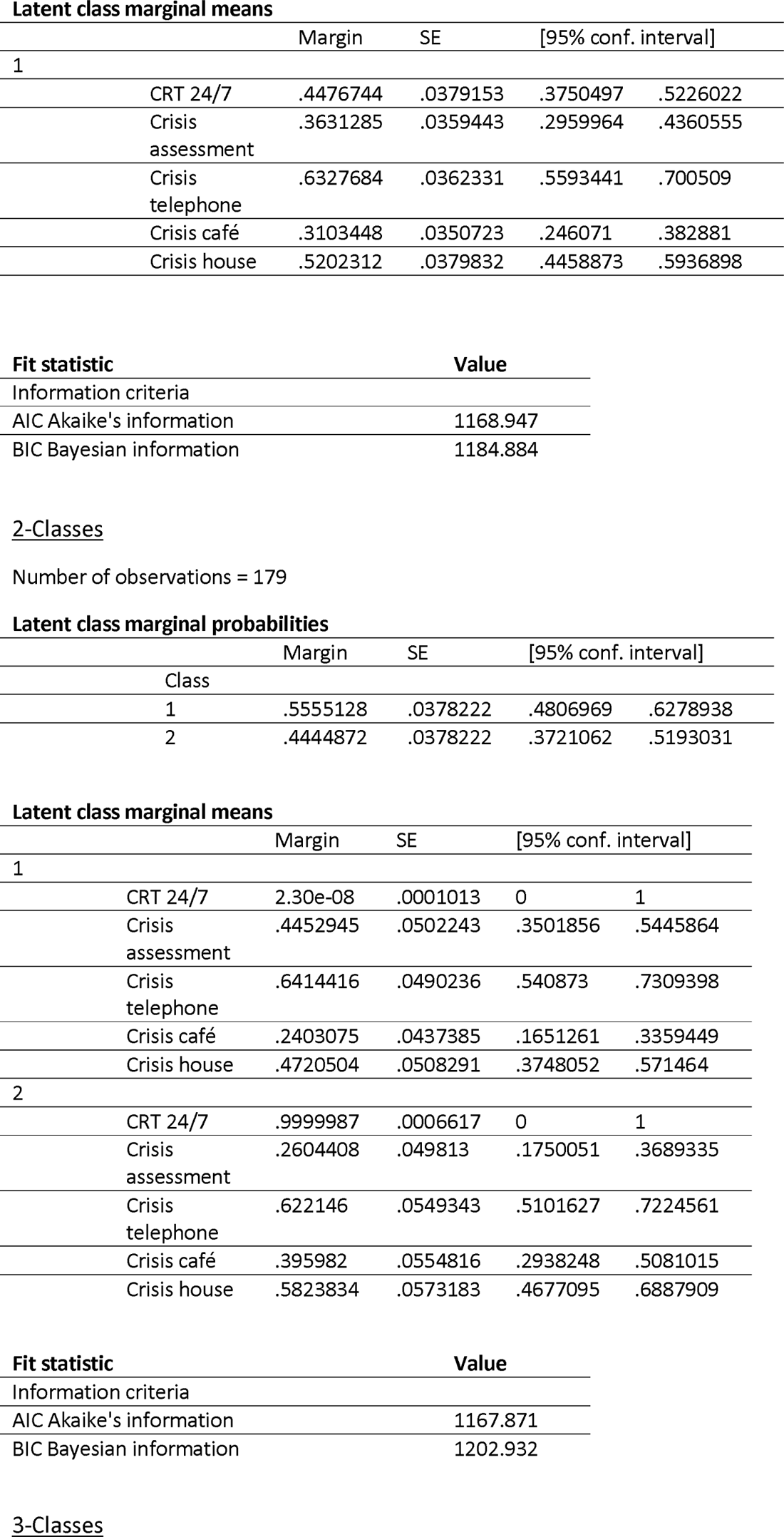
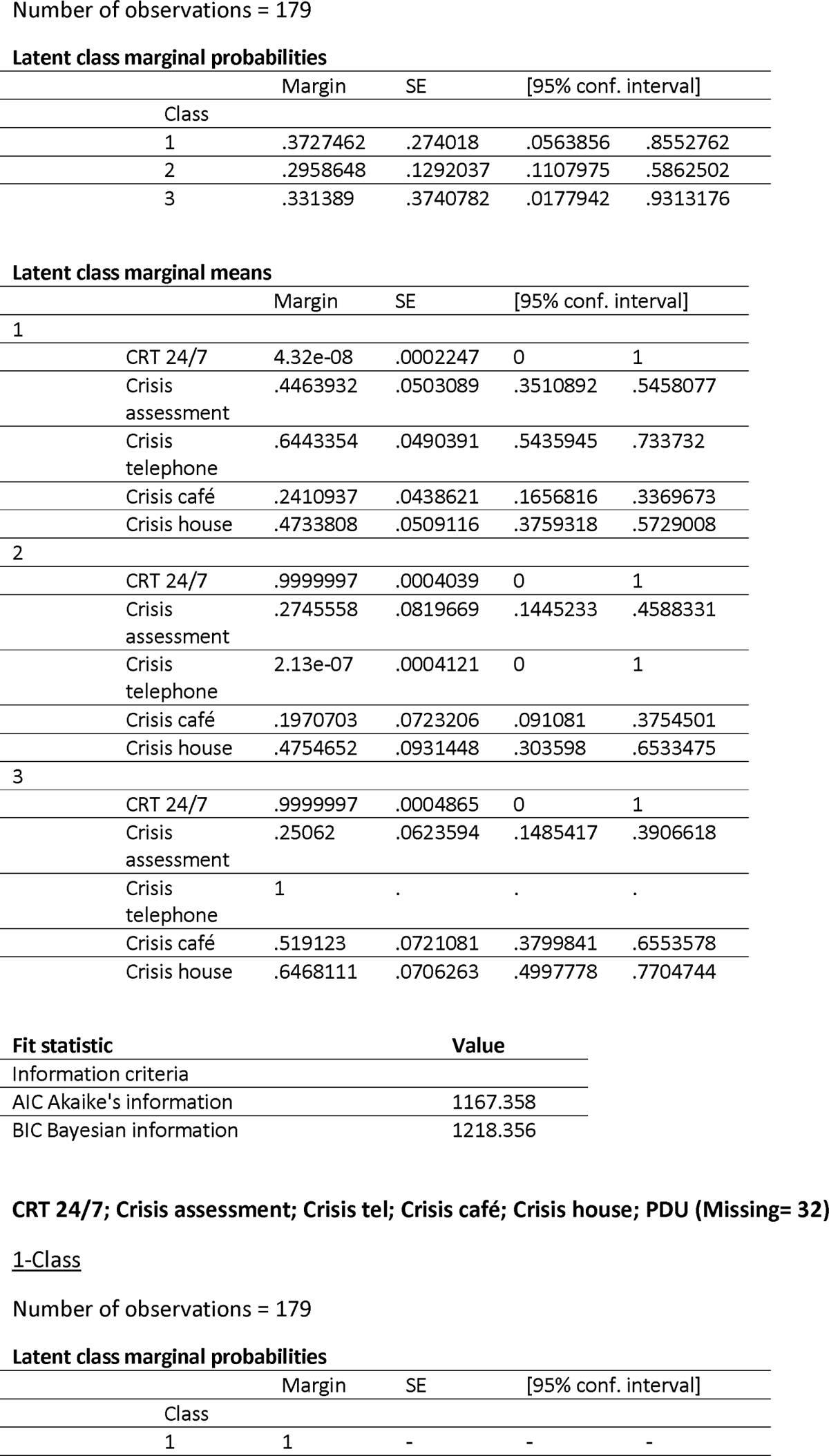
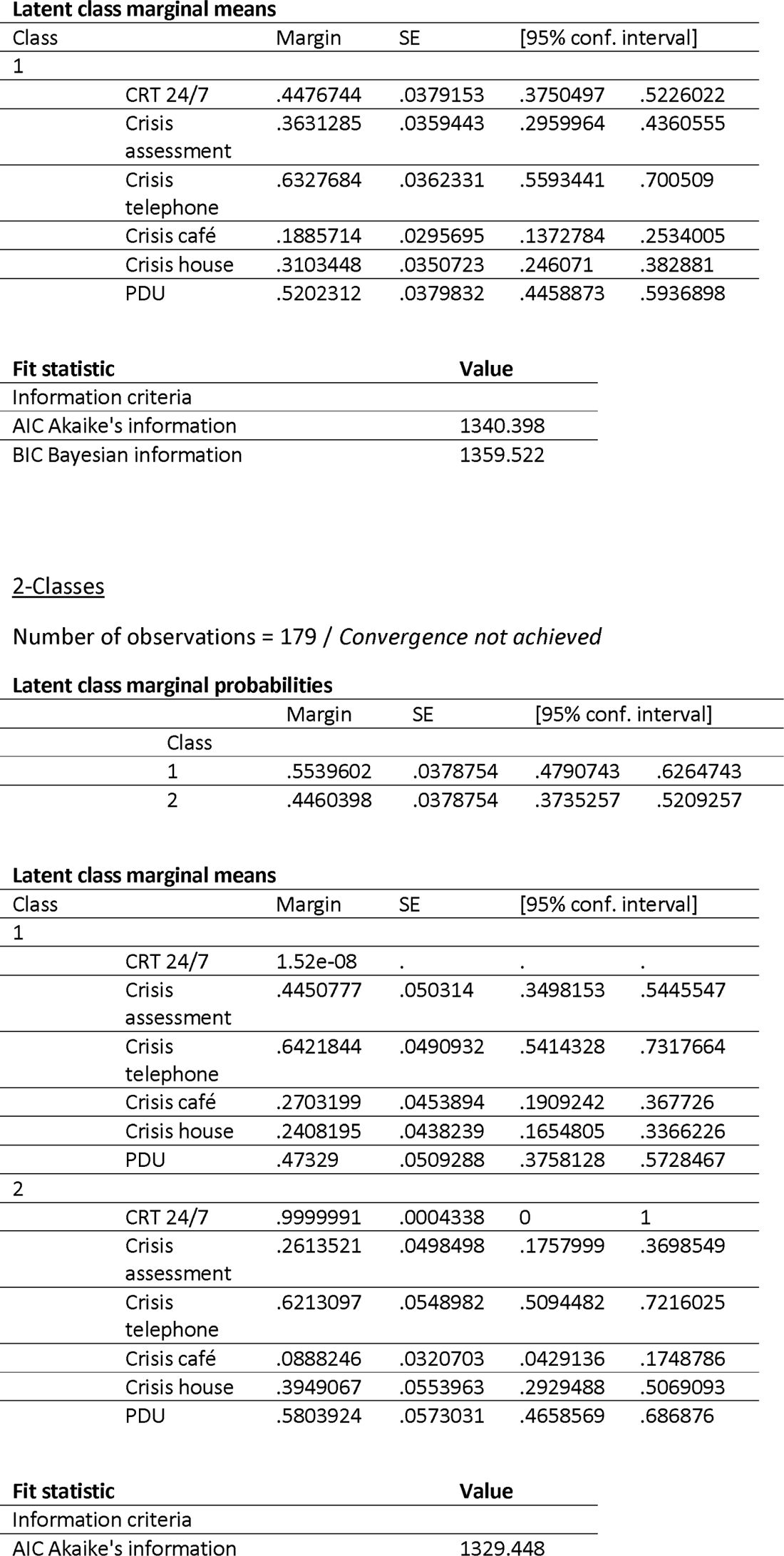
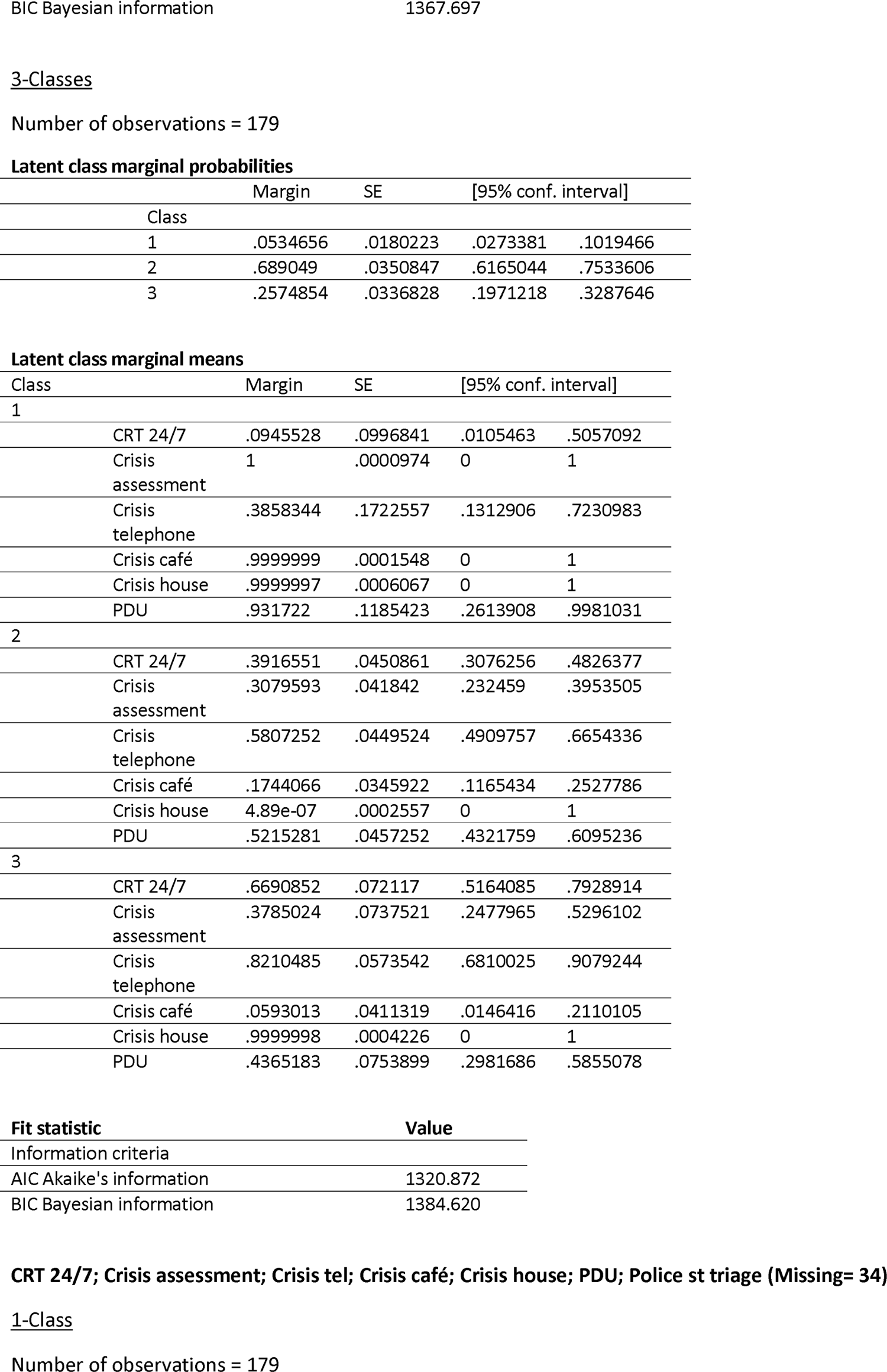
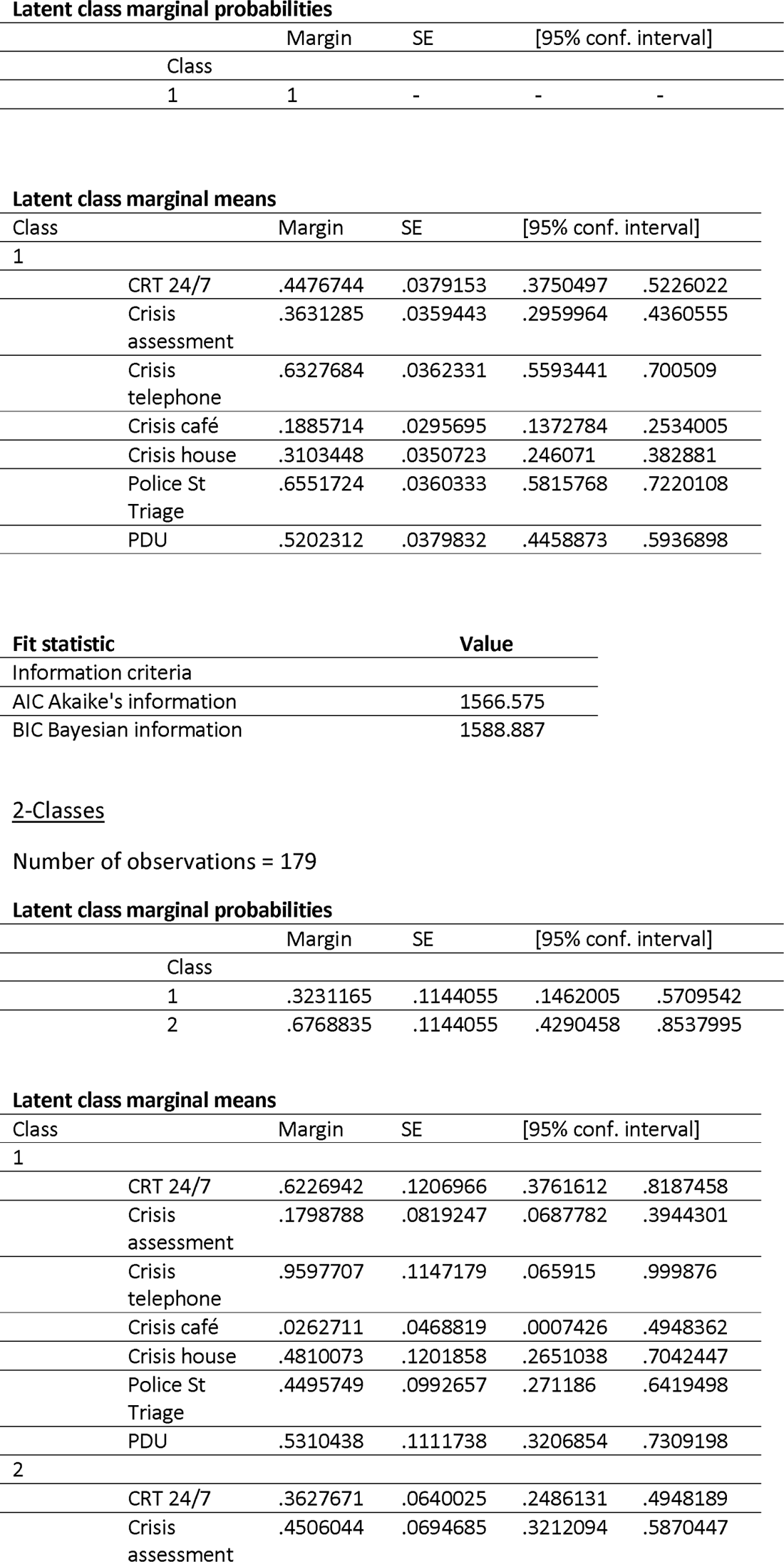
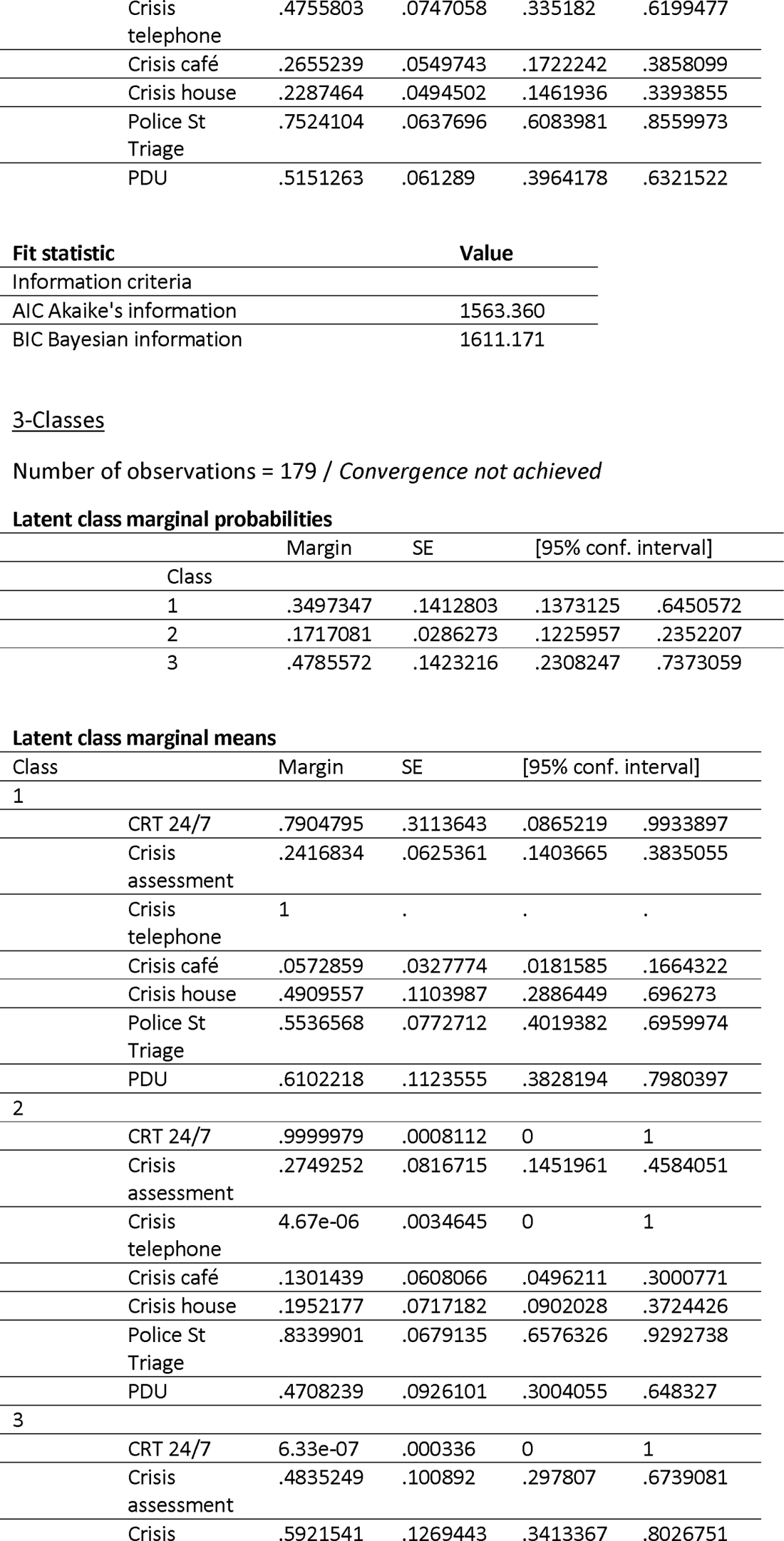
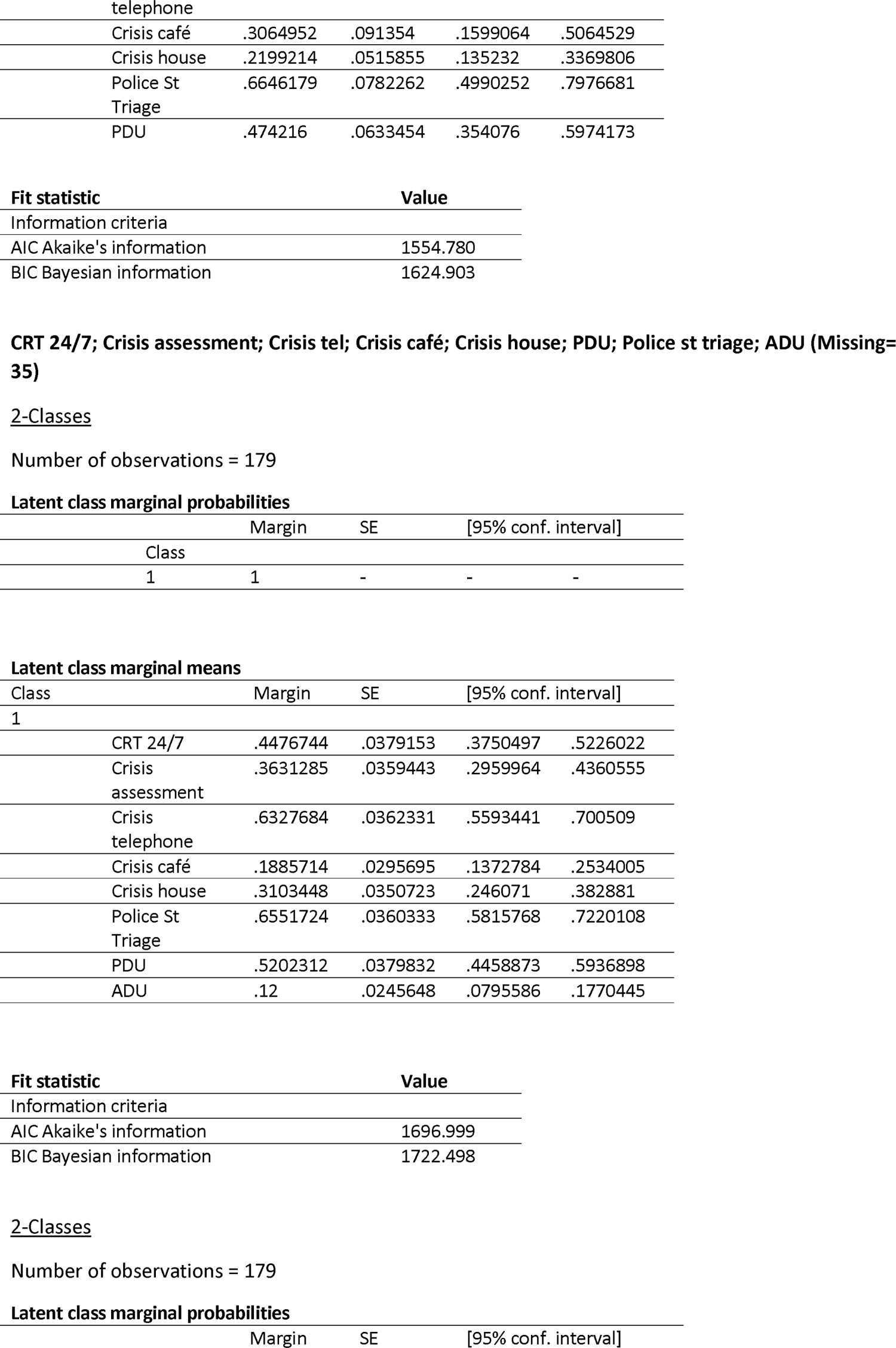
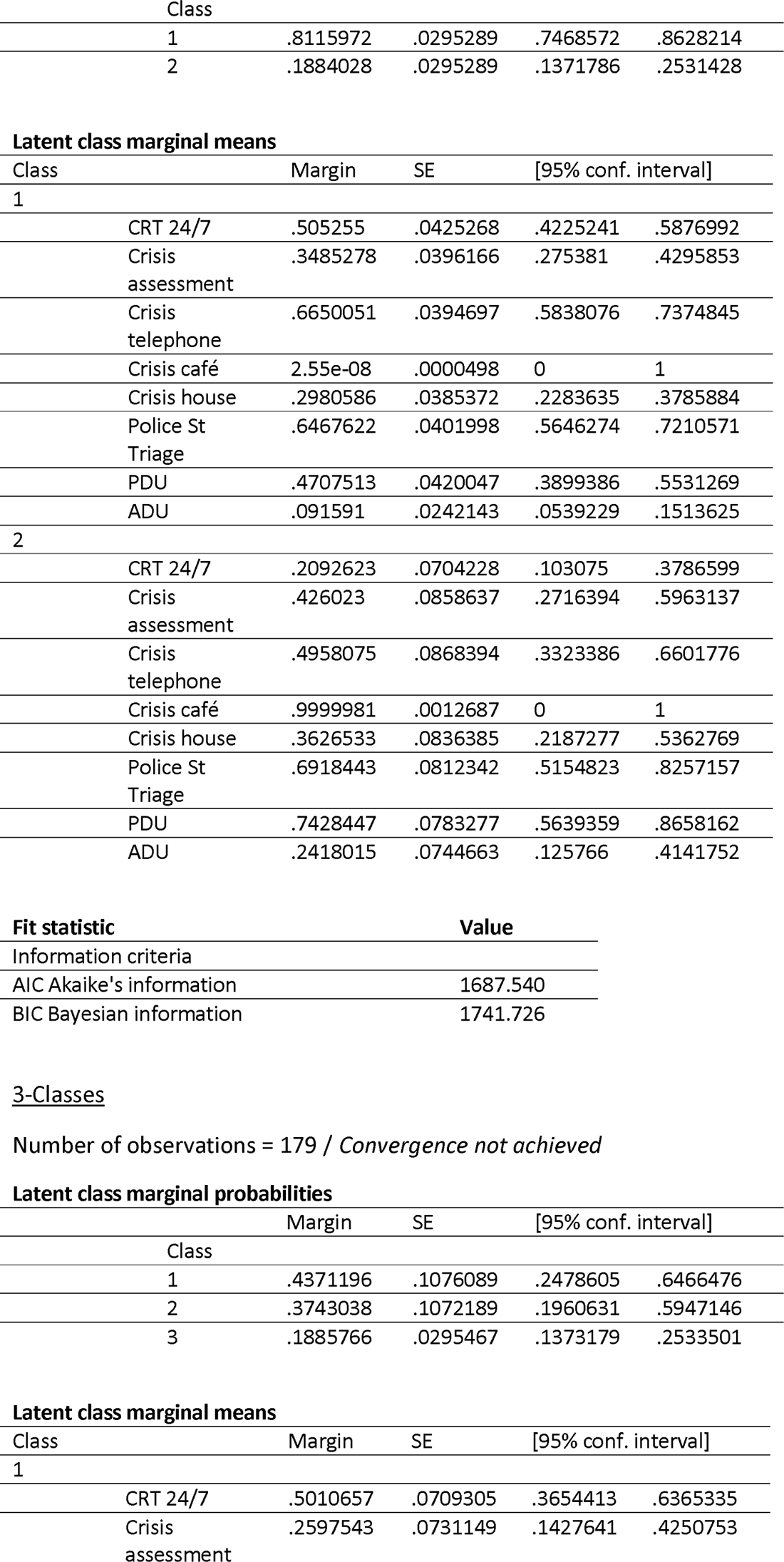
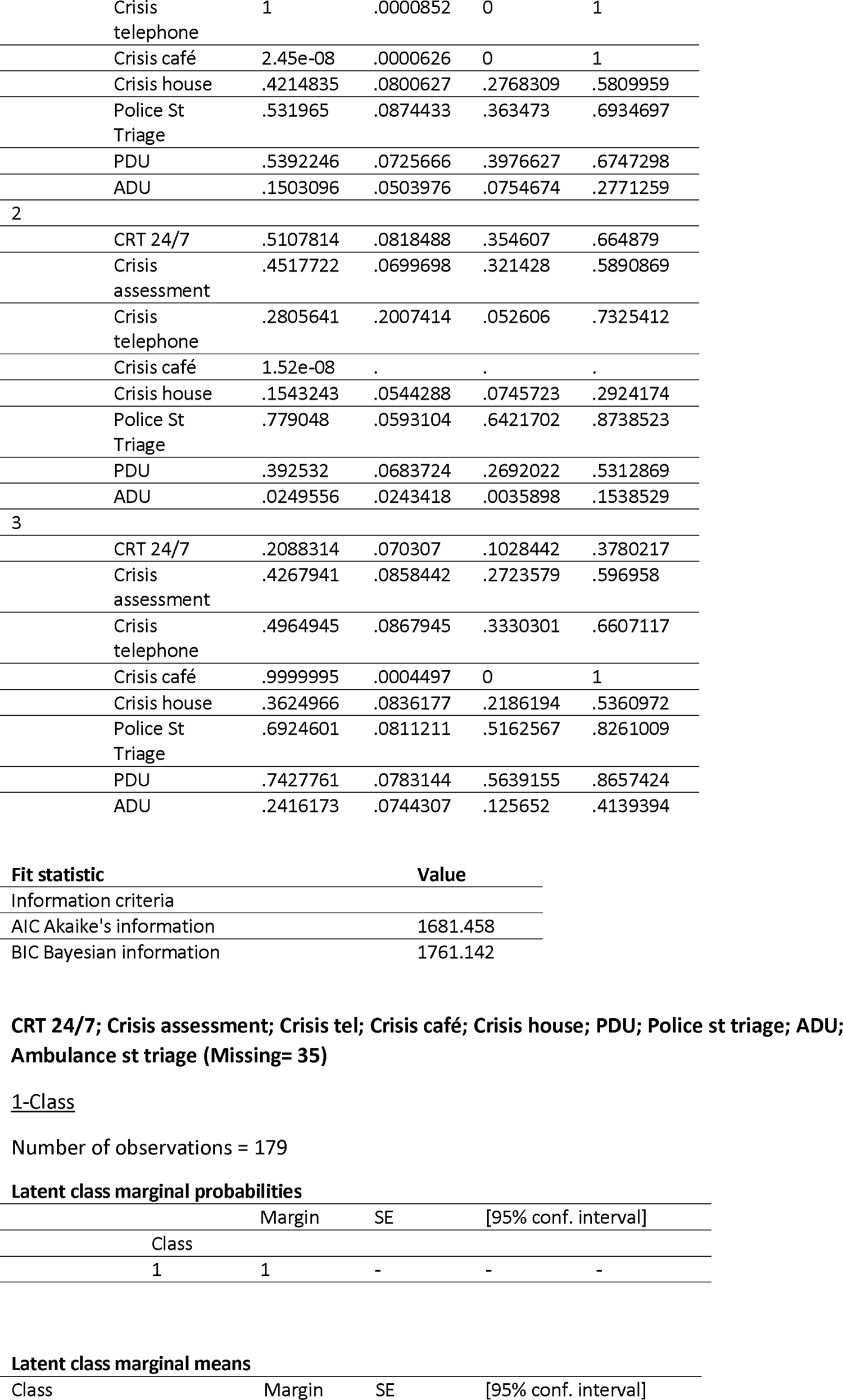
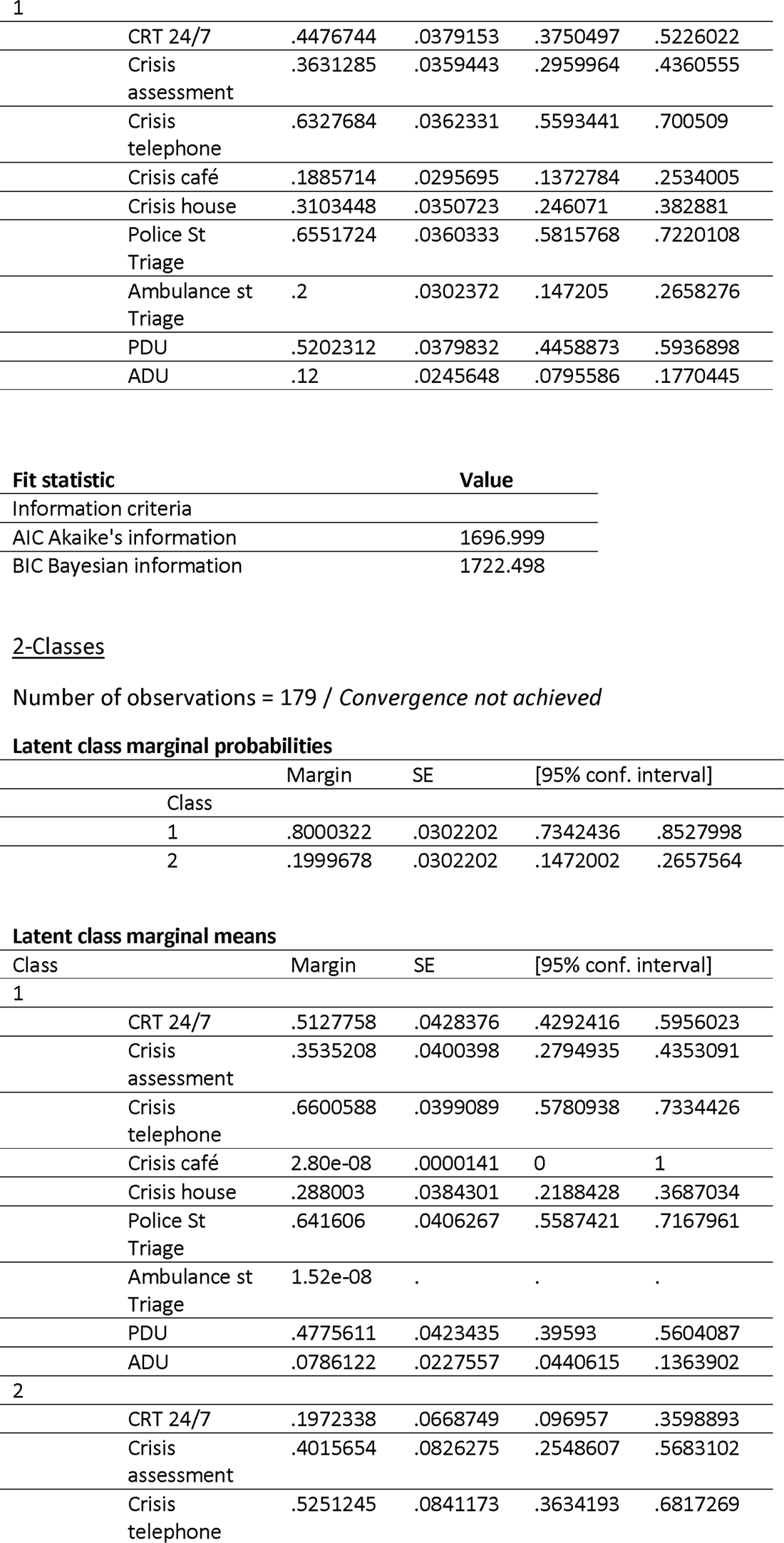
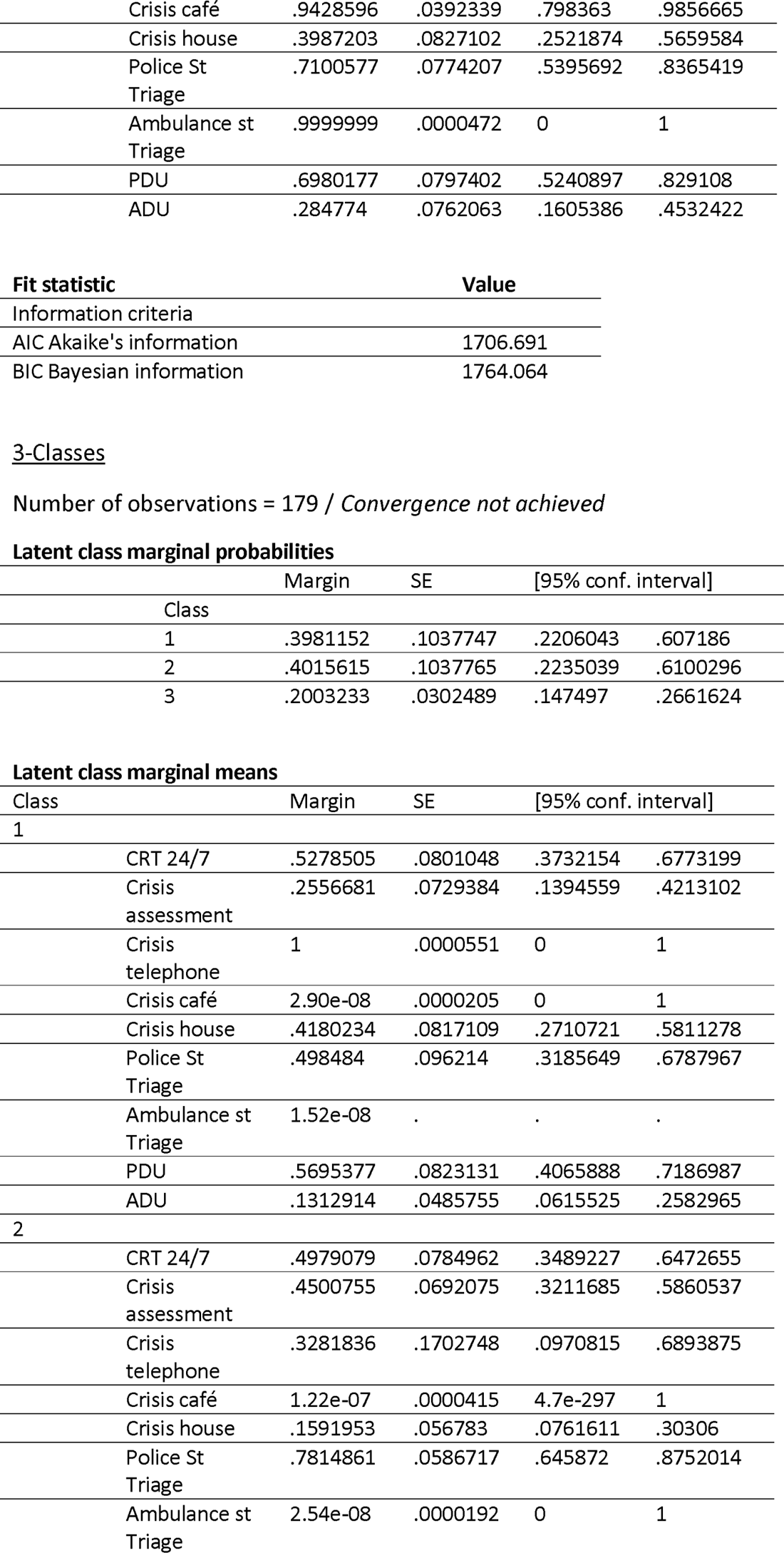
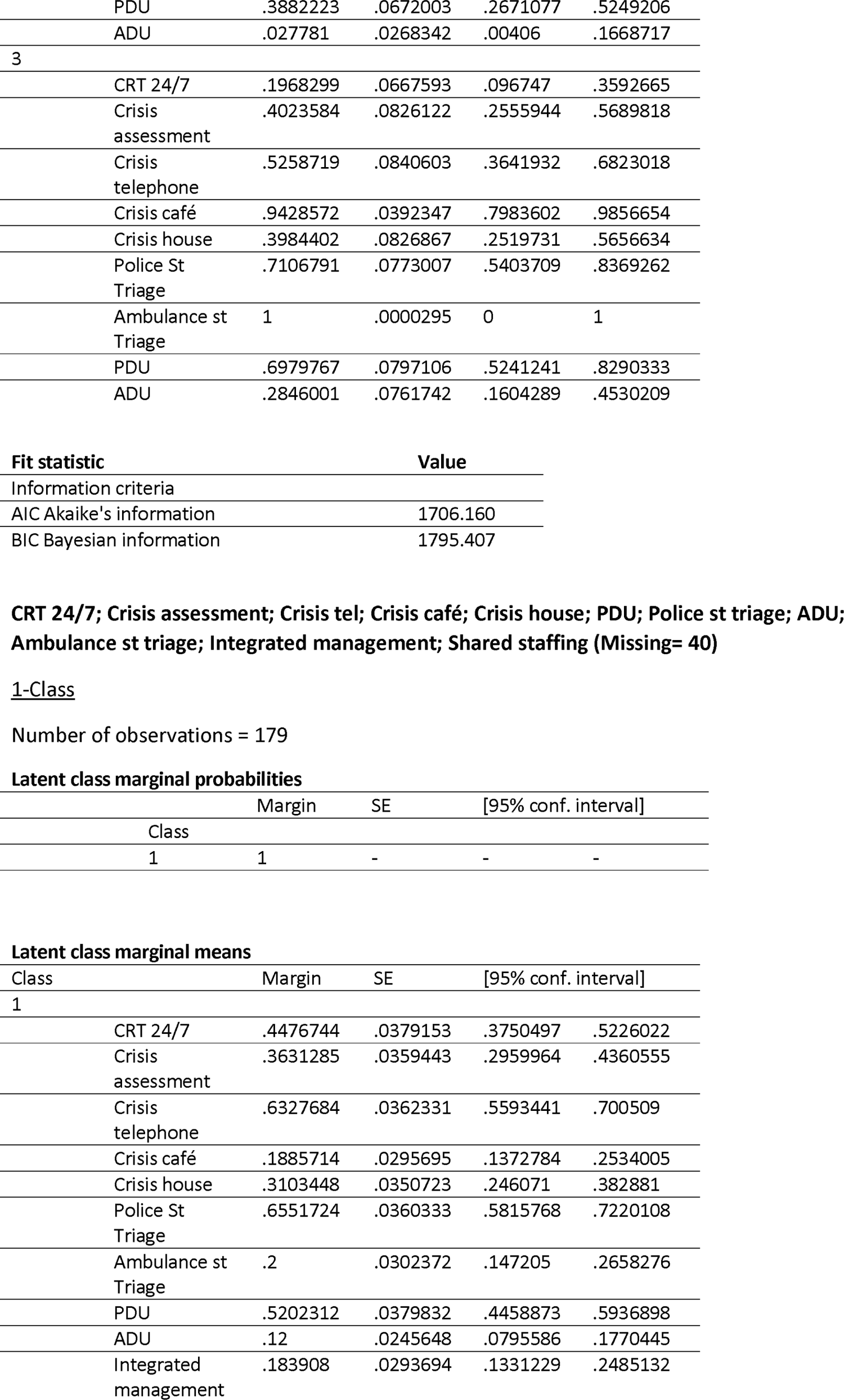
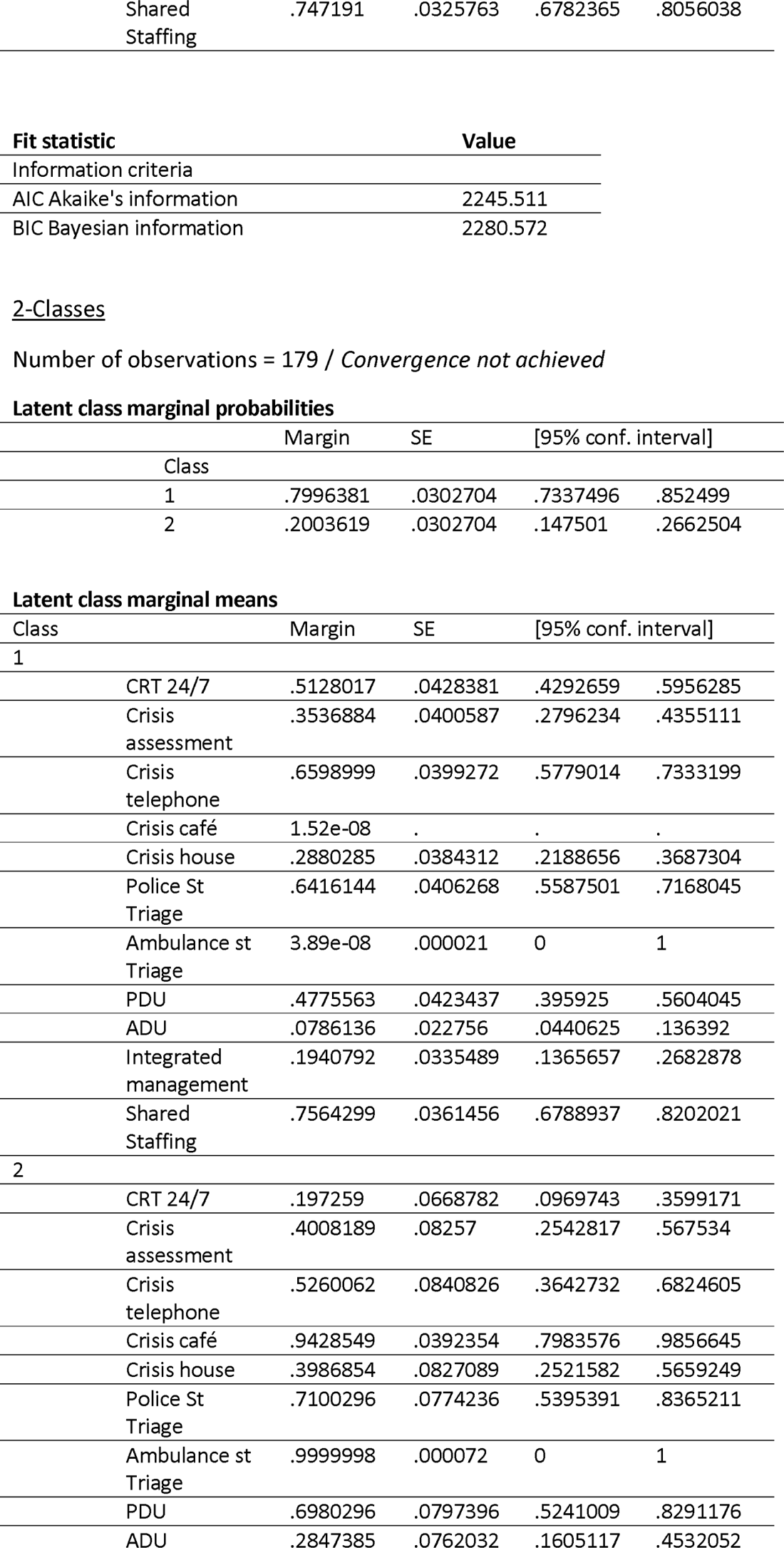
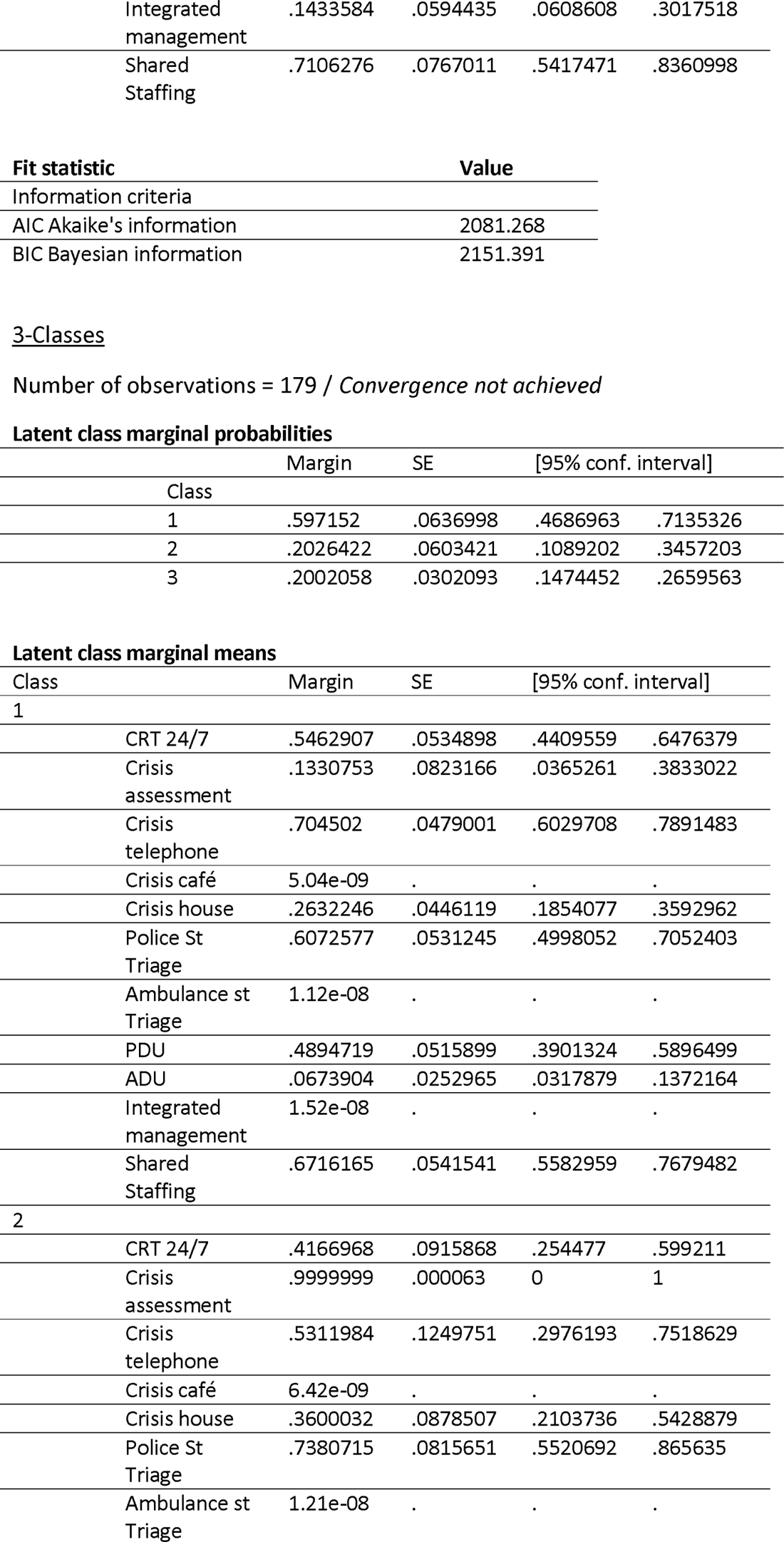
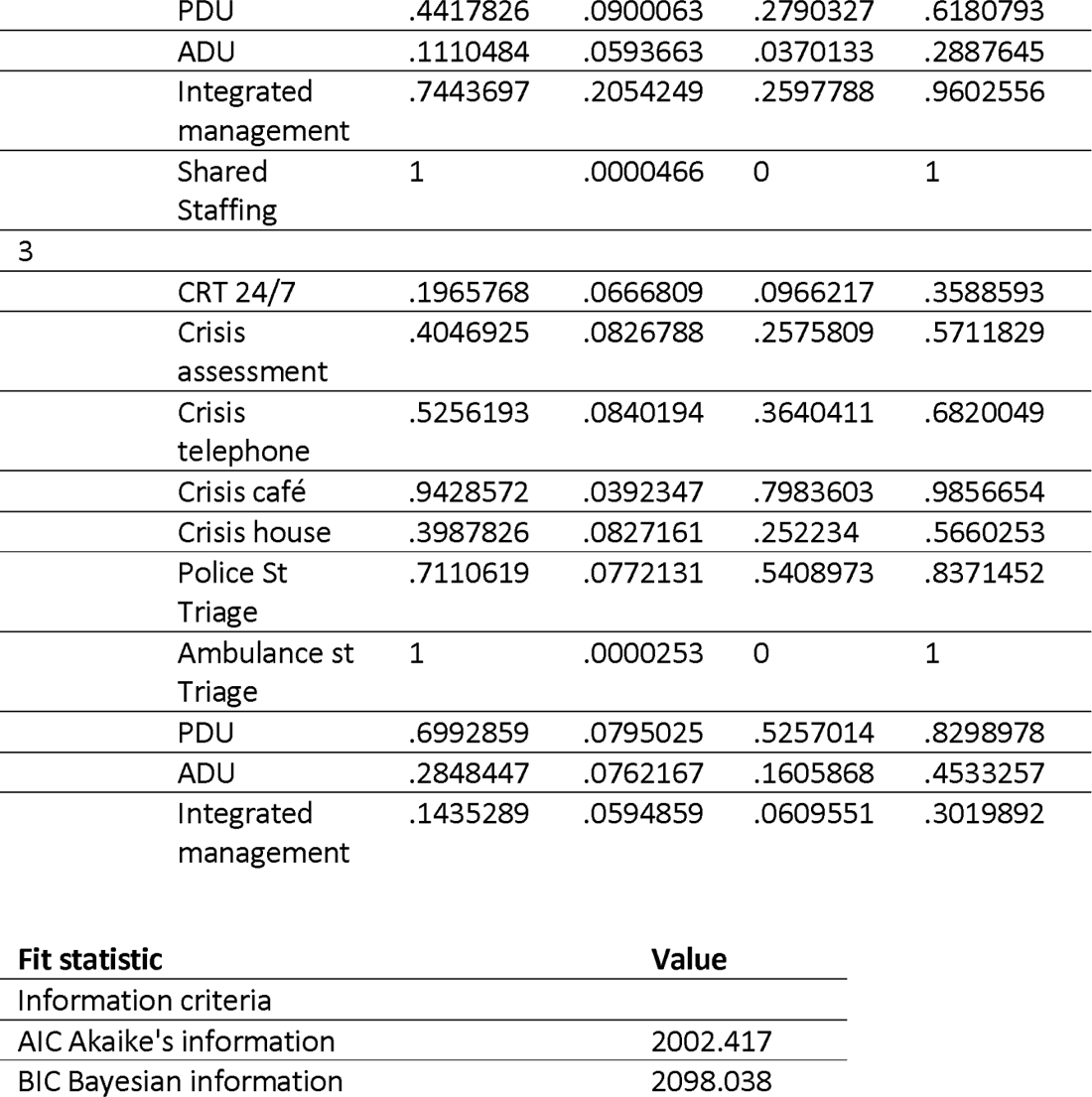

## Notes

### Competing Interest Statement

The authors have declared no competing interest.

